# Anti-spike antibody response to natural SARS-CoV-2 infection in the general population

**DOI:** 10.1101/2021.07.02.21259897

**Authors:** Jia Wei, Philippa C. Matthews, Nicole Stoesser, Thomas Maddox, Luke Lorenzi, Ruth Studley, John I Bell, John N Newton, Jeremy Farrar, Ian Diamond, Emma Rourke, Alison Howarth, Brian D. Marsden, Sarah Hoosdally, E Yvonne Jones, David I Stuart, Derrick W. Crook, Tim E. A. Peto, Koen B. Pouwels, A. Sarah Walker, David W. Eyre, the COVID-19 Infection Survey team

**Author notes:** Corresponding author: Dr David Eyre, Microbiology Department, John Radcliffe Hospital, Headley Way, Oxford, OX3 9DU. contribution considered equal (Koen B. Pouwels, A. Sarah Walker, David W. Eyre). See Acknowledgements for the Coronavirus Infection Survey team.

## Abstract

We estimated the duration and determinants of antibody response after SARS-CoV-2 infection in the general population using representative data from 7,256 United Kingdom COVID-19 infection survey participants who had positive swab SARS-CoV-2 PCR tests from 26-April-2020 to 14-June-2021. A latent class model classified 24% of participants as ‘non-responders’ not developing anti-spike antibodies. These seronegative non-responders were older, had higher SARS-CoV-2 cycle threshold values during infection (i.e. lower viral burden), and less frequently reported any symptoms. Among those who seroconverted, using Bayesian linear mixed models, the estimated anti-spike IgG peak level was 7.3-fold higher than the level previously associated with 50% protection against reinfection, with higher peak levels in older participants and those of non-white ethnicity. The estimated anti-spike IgG half-life was 184 days, being longer in females and those of white ethnicity. We estimated antibody levels associated with protection against reinfection likely last 1.5-2 years on average, with levels associated with protection from severe infection present for several years. These estimates could inform planning for vaccination booster strategies.

## Introduction

To June 2021, over 170 million severe acute respiratory syndrome coronavirus 2 (SARS-CoV-2) infections and over 3 million associated deaths have been reported globally^1^. However, in the months following infection, reinfection is uncommon and anti-spike SARS-CoV-2 antibodies are associated with protection^2–4^. The duration of post-infection immunity has important implications for the future of the pandemic and vaccination policy.^5^

Seroconversion to viral spike and nucleocapsid antigens usually happens within 1-3 weeks after SARS-CoV-2 infection^6–8^, with peak antibody levels achieved in 4-5 weeks^9,10^. However, 5-22% of individuals remain seronegative following infection^11–13^. Absence of seroconversion is more common following mild vs. severe disease (e.g. 22.2% vs. 2.6%, n=236^12^) and in asymptomatic vs. symptomatic individuals (11.0% vs 5.6% respectively, n=2,547^13^). However, the contribution of other factors, including viral load, has not been comprehensively assessed.

Among those who do seroconvert, data on the trajectory and duration of antibody responses to different SARS-CoV-2 antigens vary, partly reflecting assay-dependent differences even where similar viral antigens are studied^14,15^, as well as differences in the populations and disease groups investigated. Estimates for the half-life of anti-spike IgG antibodies (associated with neutralising activity^16^) vary from 36 to 244 days^15,17–23^. Similarly anti-nucleocapsid IgG half-lives have been estimated between 35 and 85 days^15,19,20,22^.

Most studies have had small to moderate sample sizes or specific sub-populations; large-scale representative population studies are limited. We used the Office for National Statistics (ONS) COVID-19 Infection Survey (CIS), a large community-based survey representative of United Kingdom’s general population, to investigate predictors of seroconversion following SARS-CoV-2 infection, identify anti-spike IgG antibody trajectories, and examine the peak and duration of IgG antibody responses, particularly considering the impact of demographic factors, cycle threshold (Ct) values (inversely related to viral load), and self-reported symptoms on post-infection antibody responses.

## Results

From 26 April 2020 to 14 June 2021, 467,450 participants had one or more throat and nose swab study results (median 10, IQR 8-12) during a median (IQR) 221 (141-251) days of follow-up. 19,588 (4.2%) participants ≥16 years were ever PCR-positive, 92 (0.5%) with a second episode >120 days after their first PCR-positive result (median 149, IQR 134-174 days later). Analysis included the 7,256/19,588 (37%) participants with at least one anti-spike IgG antibody measurement within [-90,+180] days of the start of their first infection episode, who contributed 14,552 antibody measurements (median 2, IQR 1-3, range 1-10; excluding measurements from 3 days after first vaccination) **(Figure S1)**.

The median age of these 7,256 participants was 47 (IQR 34-59) years, and 3,874 (53.4%) were female **(Table 1)**. 6,577 (90.6%) reported white ethnicity, 127 (1.8%) working in patient-facing healthcare, and 1,592 (21.9%) having a long-term health condition. Considering the minimum Ct across all positive tests in the first infection episode, the median was 27 (IQR 19-32), with 4,420 (60.9%) having Ct<30. 1,505 (20.7%) were only positive on a single gene (ORF1ab or N); 2,822 (38.9%) were Alpha (B.1.1.7)-compatible (S gene target failure). 4,190 (57.7%) reported having any symptoms, with 2,773 (38.2%) reporting classic symptoms (fever, cough, loss of smell, or loss of taste). 5,169 (71%) participants only contributed antibody measurements after their index positive date.

**Table 1.**
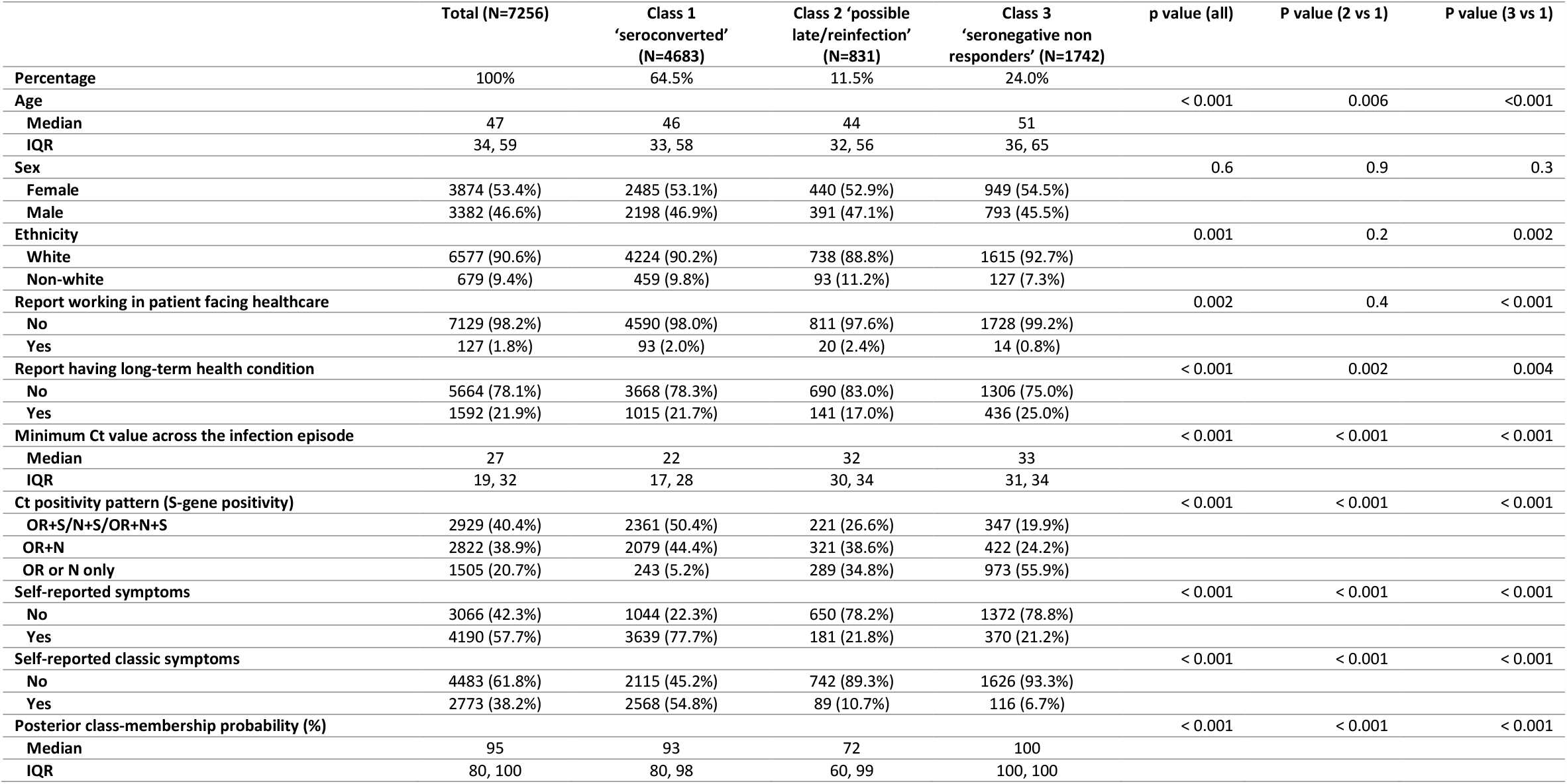
Main characteristics of participants in classes identified from latent class mixed models for 7,256 participants infected with SARS-CoV-2. Additional characteristics comparisons among classes are presented in **Table S1**, and continuous variables are presented graphically in **Figure S2**. Continuous variables were compared using Kruskall-Wallis tests, and categorical variables were compared using Chi-squared tests.

### Antibody trajectories following SARS-CoV-2 infection

A latent class analysis identified three classes of post-infection anti-spike IgG antibody responses. Class-membership probabilities were high, suggesting that participants’ responses could be reliably assigned to one of the three classes **(Figures 1&S2, Table 1;** individual trajectories shown in **Figure S3)**. Participants who seroconverted after infection comprised Class1 (n=4,683, 64.5%). These participants showed classical responses, with rises in antibody levels over the 4-5 weeks following their first PCR-positive sample, followed by subsequent waning. Class 1 had lower Ct values (median [IQR] 22 [17-28], p<0.001 vs. any other class), and a higher percentage of reported symptoms (77.7%, p<0.001 vs. any other class) and classic symptoms (54.8%, p<0.001 vs. any other class). Class 1 also had a lower percentage of single gene positives (5.2%, p<0.001 vs. any other class). 57.8% had more than one positive swab test in their first infection episode and 23.8% had a positive test in national testing programme prior to their first study positive test, a significantly higher percentage than other classes (p<0.001) **(Table S1)**.

**Figure 1.**
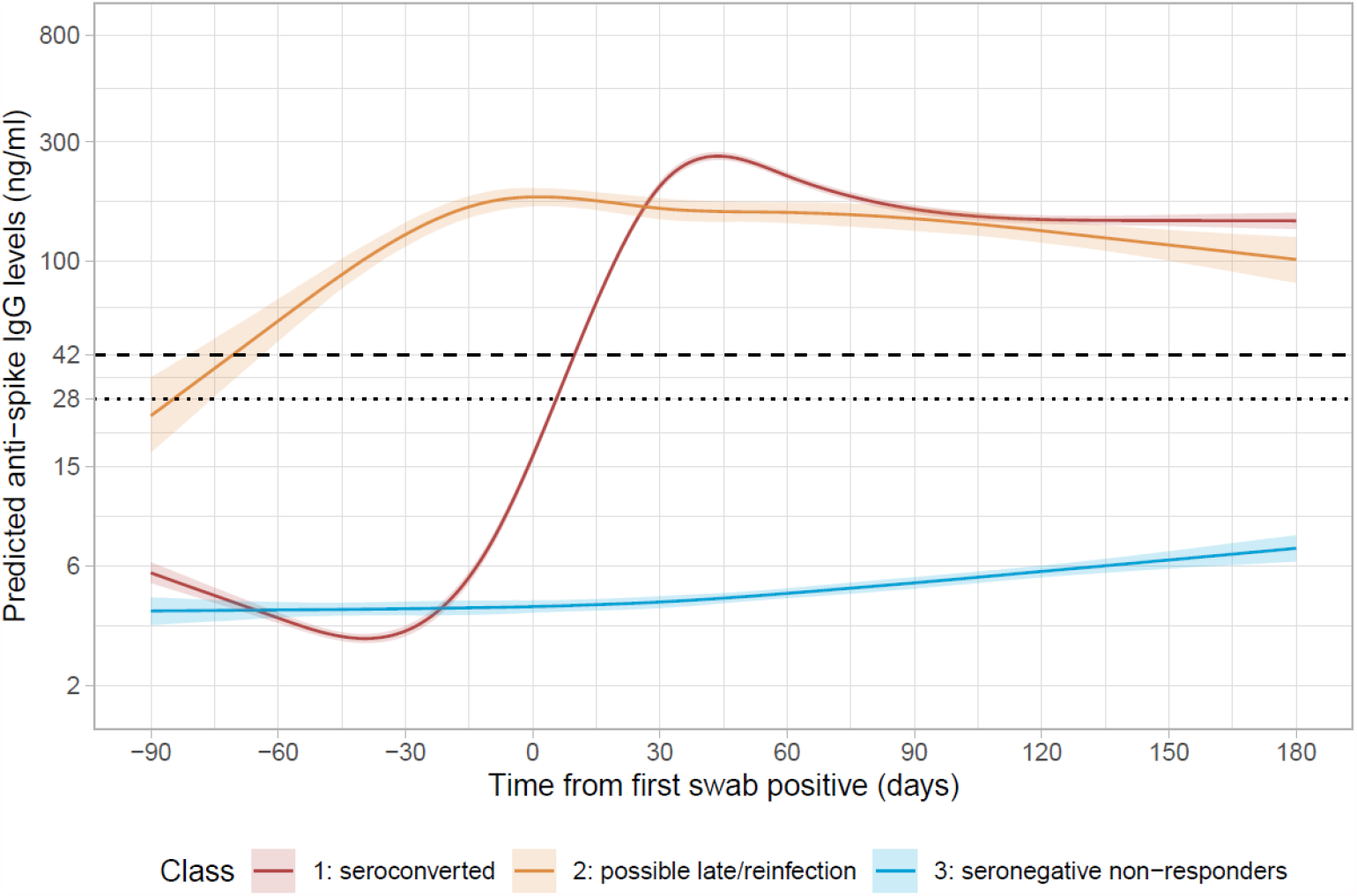
Predicted anti-spike IgG trajectories in participants with natural SARS-CoV-2 infection from latent class mixed models in 7,256 participants (with standard error of the prediction). Black dashed line indicates the assay threshold for IgG positivity (42 ng/ml) and the dotted line at 28 ng/ml (indicates level associated with 50% protection against reinfection). The 95% confidence interval are calculated by a Monte Carlo approximation of the posterior distribution of the predicted values. Restricted natural cubic splines (internal knots at -10,30,60 days, and boundary knots at -60 and 140 days) were used to model time (see methods). Distribution of the factors by class membership is shown in **Table 1**. Class 1=’seroconverted in response to infection’ (64.5%, n=4683), Class 2=’possible late/reinfection’ (11.5%,n=831), Class 3=’seronegative non-responders’ (24.0%, n=1742).

Class 2 (n=831 (11.5%), ‘possible late detection/reinfection’) also had rises in anti-spike IgG levels but these started earlier, before the index positive PCR test. Their antibody levels reached a peak around the time of the index positive and then waned. This class likely partly reflects the study design, as study PCR testing was conducted at regular, usually monthly, intervals, irrespective of symptoms, with a proportion of missed visits (see Methods). Therefore, this group could represent those where infection was detected late rather than reflecting any underlying biological difference. However, a subset may also represent reinfection with an undetected first infection. Supporting these possibilities, Ct values were higher (median [IQR] 32 [30-34]) than Class 1 (p<0.001), self-reported symptoms were less common (21.8%), as were multiple positive PCR tests (24.4%) (p<0.001) **(Tables 1&S1)**. For more participants, the index positive PCR was their first test in the study (27.4%); in the remainder, the median days since last negative was 29 days, higher than other classes (p<0.001) and with considerable skew, with 369 (44.4%) being >31 days and 256 (30.8%) being >59 days, supporting late detection contributing to this group.

Lastly, 1,742 (24.0%) participants were assigned to Class 3 (‘seronegative; non-responders’). Their IgG levels barely increased and were below the positivity threshold throughout (excepting 17 outlier individuals who appeared to mount a response >30 days after their index positive PCR test). Compared with Class 1, Class 3 had higher Ct values (median [IQR] 33 [31-34], p<0.001), a lower percentage self-reporting symptoms (21.2%, p<0.001) or classic symptoms (6.7%, p<0.001) **(Table 1)**. Very few had more than one positive swab in their first infection episode (3.4%) or an accompanying positive test in the national testing programme (1.9%) **(Table S1)**. Whilst this class would be expected to be enriched for false-positives, of 1,742 participants in this class, 595 (34%) still had strong evidence for a true-positive PCR result (Ct ≤32 and ≥2 genes detected). Class 3 were also older **(Figure S2)**, with fewer patient-facing healthcare workers (0.8%) and more participants with long-term health conditions (25.0%).

### Predictors of non-response

In the multinomial logistic regression model, independent predictors of remaining seronegative (Class 3) vs seroconverting (Class 1) were higher minimum Ct (i.e. lower viral load), not self-reporting symptoms, older age and not working in patient-facing healthcare **(Figure 2, Table S2)**, with no evidence of independent effects of sex, ethnicity, or long-term health conditions. For example, at the median age of 47 years (not working in patient-facing healthcare), the Ct threshold at which seroconversion rates reached >90% were 26, 23 and 17 for those reporting classic symptoms, other symptoms or no symptoms (**Figure 2b**). Excluding Ct from the model, there was still no evidence of independent effects of long-term health conditions, but non-white ethnicity was associated with lower odds of being in Class 3 (OR=0.70, 95%CI 0.55-0.90, p=0.005) than Class 1.

**Figure 2.**
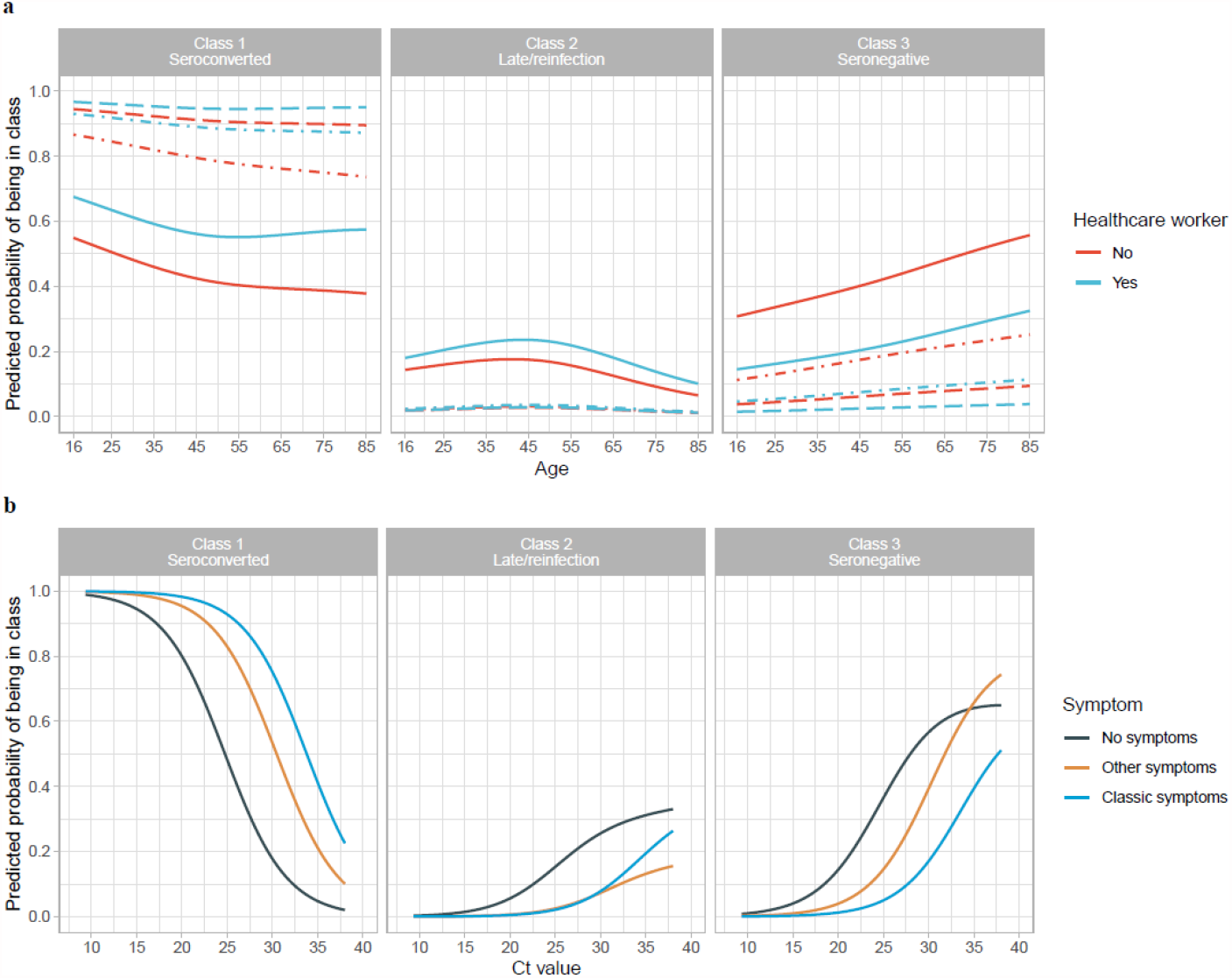
Predicted probability of being in Class 1 (seroconverted in response to infection), 2 (’possible late/reinfection), and 3 (seronegative non-responders). **a**, By age and working in patient-facing healthcare, plotted at the reference category for other variables (female, white ethnicity, no long-term health condition, Ct=26, have only one positive swab test during the infection episode) and no symptoms (solid line), other symptoms (dash-dotted line), classic symptoms (dashed line). **b**, By Ct value and self-reported symptoms, plotted at the reference category for other variables (age=47y, female, white ethnicity, no long-term health condition, not working in patient-facing healthcare, have only one positive swab test during the episode). Age was fitted using natural cubic spline with one internal knot placed at 50 years and two boundary knots at 20, 80 years. Full model results are shown in **Table S2**.

**Figure 3.**
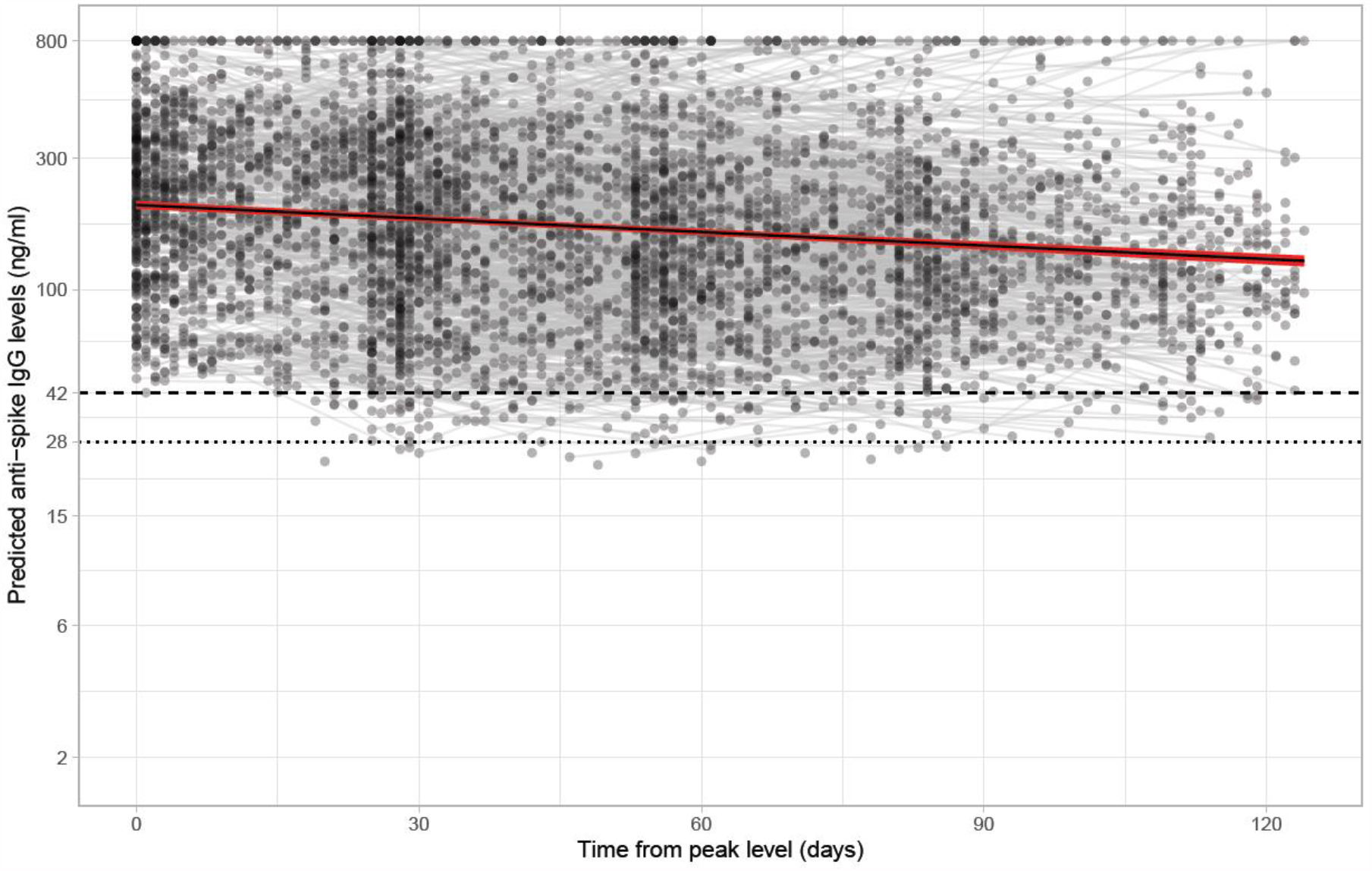
Estimated mean trajectory of anti-spike IgG antibody levels and individual trajectories in 3,271 participants in Class 1. The timing of the peak level 56 days after the first positive swab is determined from the latent class mixed model. The posterior mean and 95% credibility interval are shown with the black line and red shaded area. Black dashed line indicates the assay threshold for IgG positivity (42 ng/ml) and the dotted line at 28 ng/ml indicates level associated with 50% protection against reinfection.

To investigate associations with specific symptoms, we fitted a logistic regression model comparing only seroconversion (Class 1) vs non-response (Class 3), and omitting Ct and other test characteristics as these may mediate effects of symptoms. We found cough, loss of smell, fever, loss of taste, fatigue, headache, and sore throat were associated with lower odds of non-response, with cough (OR=0.20, 95%CI 0.15-0.25, p<0.001) and loss of smell (OR=0.21, 95%CI 0.13-0.33, p<0.001) mostly strongly associated. Results remained similar restricting seronegatives to those with stronger evidence of a true PCR-positive result (Ct ≤32 and ≥2 genes detected) **(Table 2, Figure S4)**. We additionally examined the association with specific comorbidities by incorporating them into the model but found no strong evidence of major impact **(Table S3)**.

**Table 2.** Odds ratio with 95% confidence intervals from logistic regression comparing seronegative vs seroconverting (Class 3 vs Class 1) using demographic factors and individual symptoms that would be available without a positive test result. (A) Using all data from Class 3 (N=1,742) (B) Restricting Class 3 to those with Ct value ≤32 and ≥2 genes detected (N=595) to decrease the impact of potential false positive swab tests. Age was fitted using natural cubic spline with one internal knot placed at 50 years and two boundary knots at 20, 80 years. Effect of age is presented in **Figure S4**. The 95% confidence intervals are calculated by prediction ± 1.96*standard error of the prediction; Wald p values are shown.

|  | Class 1<br>'seroconverted' | Class 3 (A: Using all data):<br>'seronegative non responders' |  |  | Class 3 (B: Conditioning on Ct ≤32<br>and ≥2 gene positivity) |  |  |
| --- | --- | --- | --- | --- | --- | --- | --- |
|  |  | OR | 95%CI | p-value | OR | 95%CI | p-value |
| Age | ref |  |  | **<0.001** |  |  | **<0.001** |
| Sex (Male vs Female) |  | 0.84 | 0.75-0.96 | **0.008** | 0.87 | 0.73-1.04 | 0.1 |
| Ethnicity (Non-White vs White) |  | 0.75 | 0.60-0.94 | **0.01** | 0.88 | 0.63-1.20 | 0.4 |
| Report having long-term health conditions (Yes vs No) |  | 1.15 | 0.99-1.34 | 0.06 | 0.97 | 0.78-1.21 | 0.8 |
| Report working in patient facing healthcare (Yes vs No) |  | 0.42 | 0.22-0.75 | **0.005** | 0.44 | 0.15-1.02 | 0.09 |
| Cough (Yes vs No) |  | 0.20 | 0.15-0.25 | **<0.001** | 0.24 | 0.16-0.35 | **<0.001** |
| Loss of smell (Yes vs No) |  | 0.21 | 0.13-0.33 | **<0.001** | 0.34 | 0.17-0.62 | **<0.001** |
| Fever (Yes vs No) |  | 0.42 | 0.29-0.60 | **<0.001** | 0.51 | 0.29-0.84 | **0.01** |
| Loss of taste (Yes vs No) |  | 0.53 | 0.35-0.79 | **0.002** | 0.51 | 0.27-0.89 | **0.03** |
| Fatigue/weakness (Yes vs No) |  | 0.58 | 0.44-0.75 | **<0.001** | 0.69 | 0.46-1.01 | **0.06** |
| Headache (Yes vs No) |  | 0.69 | 0.54-0.87 | **0.003** | 0.54 | 0.37-0.79 | **0.002** |
| Sore throat (Yes vs No) |  | 0.69 | 0.52-0.91 | **0.01** | 0.59 | 0.37-0.91 | **0.02** |
| Myalgia (Yes vs No) |  | 0.75 | 0.55-1.02 | 0.07 | 0.70 | 0.43-1.09 | 0.1 |
| Nausea/Vomiting (Yes vs No) |  | 0.93 | 0.59-1.42 | 0.7 | 1.25 | 0.66-2.25 | 0.5 |
| Diarrhoea (Yes vs No) |  | 1.06 | 0.66-1.68 | 0.8 | 0.88 | 0.39-1.79 | 0.7 |
| Abdominal pain (Yes vs No) |  | 1.07 | 0.62-1.78 | 0.8 | 0.98 | 0.41-2.07 | 1 |
| Shortness of breath (Yes vs No) |  | 1.39 | 0.97-1.95 | 0.07 | 1.60 | 0.97-2.57 | 0.06 |

### Determinants of the peak and half-life of antibody responses

We estimated anti-spike IgG peak antibody levels and half-life post-infection using participants predicted to belong to Class 1, i.e. showing a classical antibody response, excluding those in Class 2 where the timing of first infection was unclear and those who remained seronegative in Class 3. We estimated trajectories from 56 days after the first positive in the infection episode, when the IgG levels were close to the maximum level with high data completeness **(Figure S5)**. 3,271 participants were included in this analysis, contributing 5,148 antibody measurements (interval censored at an assay upper limit of 800 ng/ml mAb45 equivalent units), median (IQR) [range] 1 (1-2) [1-5] per participant. Using a Bayesian linear mixed model, assuming antibody levels fell exponentially (i.e. linearly on the log scale) and accounting for variation in individuals’ peak levels and half-lives using correlated random effects, the estimated mean anti-spike IgG half-life was 184 days (95% credibility interval, Crl 163-210), and peak level was 203 ng/ml (95%Crl 190-210) **(Figure 2)**. Estimated peak levels varied substantially between participants, ranging from 42 to 1,390 ng/ml **(Figure S6a)**. Longer half-lives were correlated with lower peak levels **(Figure S6b)** (Spearman’s rank coefficient=-0.50, p<0.0001; correlation between random intercept and slope -0.26). Results were similar in sensitivity analyses starting modelling from different times and using different interval censoring thresholds (400, 500 ng/ml) **(Table S4)**.

In the multivariable linear mixed model, age, ethnicity, and Ct values were independently associated with IgG peak levels (model intercept), while sex and ethnicity were independently associated with IgG half-life (model slope) **(Table 3, S5, Figure S7;** posterior checks and MCMC diagnostics in **Table S5, Figures S8, S9)**. Conditional on having seroconverted (which occurred at lower rates in older individuals), older age was associated with higher IgG peak levels (adjusted 18 ng/ml higher (95%Crl 13-23) per 10 years older). Males had a shorter half-life than females (adjusted 77 days shorter, 95%Crl 23-178). Non-white participants had higher IgG peak levels (adjusted 82 ng/ml higher (95%Crl 55-113) than white participants, but a shorter half-life (adjusted 75 days shorter, 95% Crl 1-181). Higher Ct values (i.e. lower viral burden) were associated with a slightly higher peak level (adjusted 1 ng/ml higher (95% Crl 0-2) per 1 unit higher). Conditional on inclusion in the analysis, i.e. seroconversion, we did not find any evidence of effect of reported long-term health conditions or self-reported symptoms on either IgG peak levels or half-life.

**Table 3.**
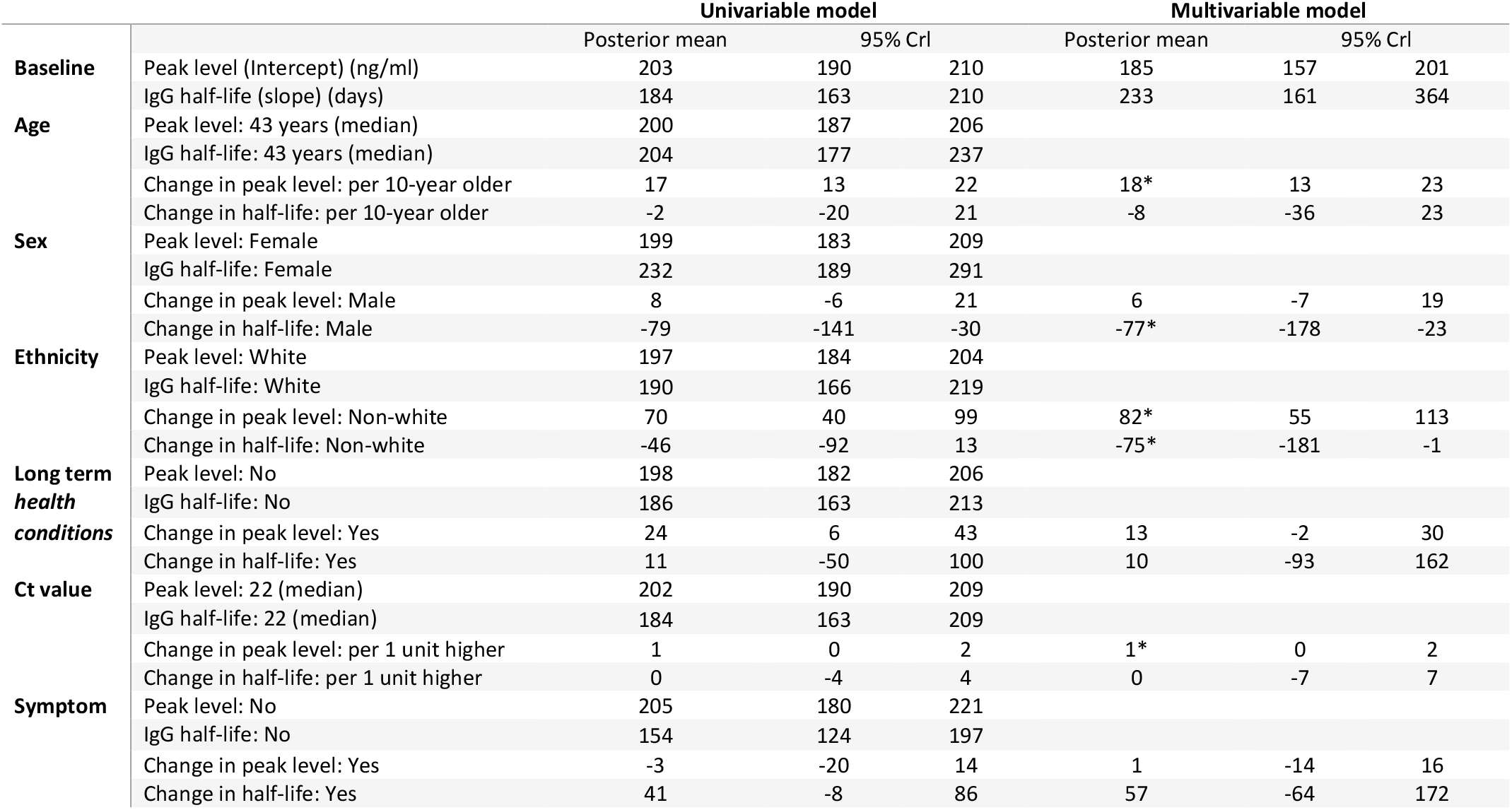
Posterior mean and 95% credibility intervals for anti-spike IgG peak level (intercept) (ng/ml) and half-life (slope) (days) in the univariable and multivariable models in 3,271 participants in Class 1. The reference category in the multivariable model is: 43-year-old, female, white ethnicity, no long-term health conditions, Ct value=22, and no self-reported symptoms. * indicates where the multivariable 95% CrI excludes 0 (no effect).

### Duration of antibody responses and possible associated immune protection

Using the multivariable linear mixed model, we estimated that antibody responses were likely to remain positive, i.e. ≥42ng/ml, for 552 (95%Crl 316-989), 408 (251-623), 476 (306-798), 385 (258-565) days from the start of infection for white females, white males, non-white females, non-white males aged 60 years, respectively. From the start of infection to 28ng/ml, the antibody level associated with 50% protection against new infection in a study of those previously infected^4^, the estimated time was 673 (418-1,196), 495 (330-737), 562 (378-917), 448 (317-651) days respectively. For a threshold of 6ng/ml, estimated to provide 50% protection against severe infection (based on previous estimates that this was provided by neutralising antibody levels 3% of peak^24^), the estimated time was 1,140 (781-1,966), 819 (605-1,158), 884 (628-1,448), 699 (525-1,000) days respectively (**Figure 4, Table S6**).

**Figure 4.**
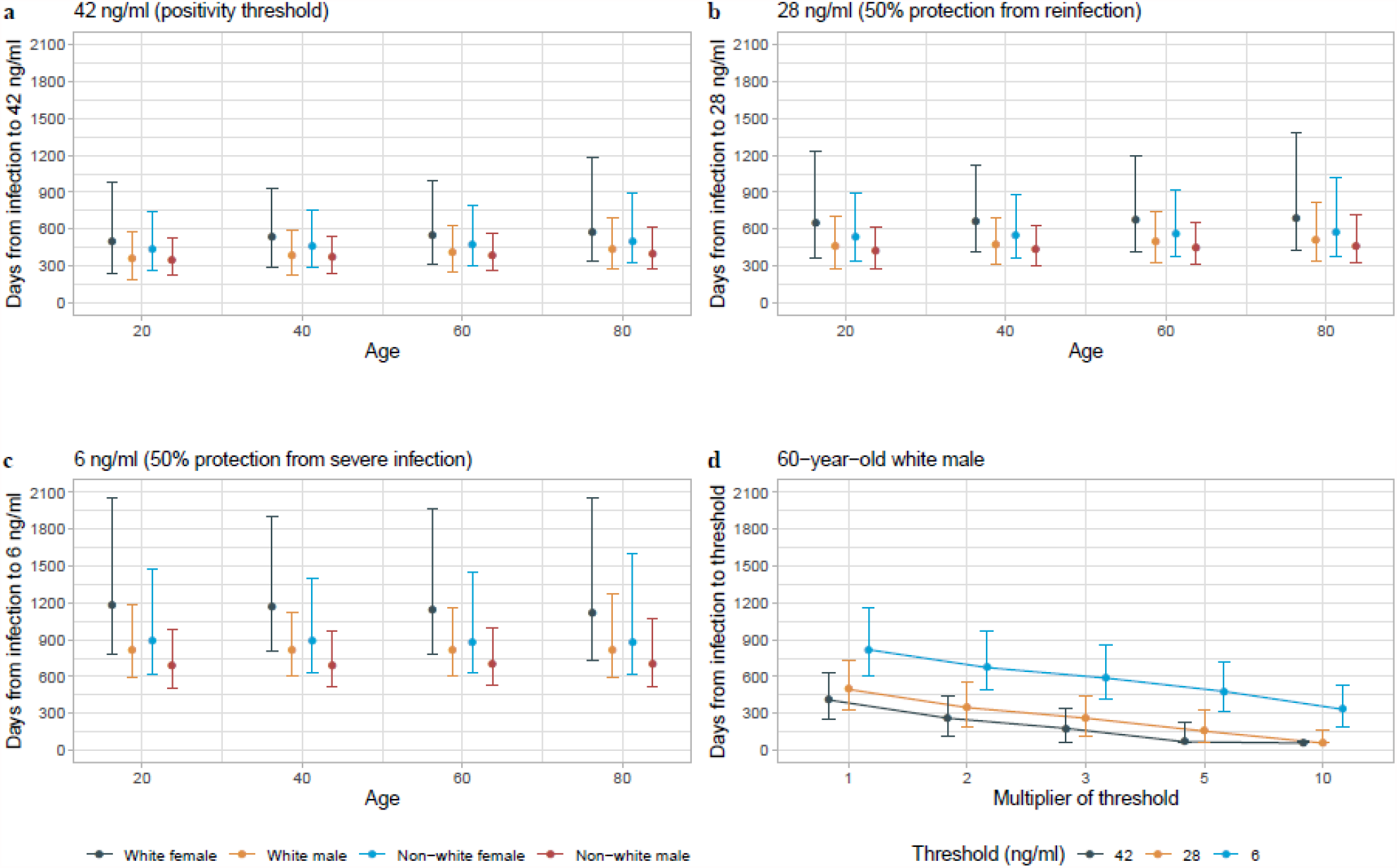
Posterior predicted time (95% credibility interval) the start of infection to three anti-spike IgG thresholds (42, 28, and 6 ng/ml) by age (20, 40, 60, 80 years), sex, and ethnicity from the multivariable model. **a**, Time from the start of infection to the positivity threshold of 42 ng/ml. **b**, Time from the start of infection to the equivocal threshold of 28 ng/ml, which corresponds to 50% protection against PCR-confirmed reinfection. **c**, Time from the start of infection to 6 ng/ml, which corresponds to 50% protection against severe infection. **d**, Time from the start of infection to the above three thresholds multiplied by 2, 3, 5, and 10, in a 60-year-old white male as an example, to estimate the duration given the higher antibody level required for protection against variants of concern. Estimations are shown in **Table S6**.

To allow for emerging viral variants needing higher antibody concentrations to afford the same level of neutralising activity, a sensitivity analysis assumed 1-to 10-fold greater antibody concentrations were required. For example, if 5-fold higher concentrations were required, for an example 60-year-old white male, the estimated duration of response was 68 (57-222), 155 (57-323), and 479 (312-714) days for levels associated with a positive result (5*42ng/ml), 50% protection from infection (5*28ng/ml), and 50% protection against severe infection (5*6ng/ml) respectively.

## Discussion

We use data from a representative national UK survey to determine predictors of seroconversion following a positive PCR test and investigate the duration of antibody responses and possible associated protection in those who do seroconvert.

We found 24% of participants did not seroconvert after testing PCR-positive, including 34% of participants with strong evidence for a true positive PCR result (Ct≤32, ≥2 genes detected). Similar observations have been reported before, but with varying percentages of non-responders from 0%-25%^11–13,25–28^. Non-responders likely include both genuine non-responders and false-positive results. Consistent with both possibilities, non-responders had fewer symptoms and higher Ct values (lower viral loads), but, more consistent with being genuine non-responders, they were also older. We found no evidence of an independent effect of long-term health conditions on non-response, possibly reflecting the heterogeneity of this group including those with a range of cardiovascular and metabolic conditions not typically associated with impaired humoral immunity, as well as conditions more directly impacting antibody production (e.g. hypogammaglobulinaemia). Other studies have reported that people taking immunosuppressive medications or with impaired immunity have decreased antibody responses, including those with diabetes, HIV, lymphoma, inflammatory bowel disease, and those taking non-steroidal anti-inflammatory drugs^29–33^. Although in some populations, antibodies are associated with protection from reinfection^3,4^, the risk of reinfection and vaccine failure in PCR-positive seronegative individuals from specific immunocompromised groups needs further study.

Although the specificity of PCR testing in this cohort has been estimated to be at least 99.995%^34,35^, given the large number of tests performed in asymptomatic individuals, i.e. with a low pre-test probability of infection, assuming a sensitivity of 94%^36^ and specificity 99.995%, the positive predictive value of PCR tests ranges between 95.0% and 99.7% for true SARS-Cov-2 prevalences between 0.1% and 2%. The majority (96.6%) of participants in Class 3 only had one positive swab test during the study, a much higher percentage than the other Classes, and only 1.9% had positive test results from the national testing programme; however mild and asymptomatic infections would also be expected to result in only one positive swab on a monthly testing schedule and no national testing programme result.

Whether certain characteristics can predict whether people develop antibodies or not following a positive PCR test is of great interest to the public. We found that apart from age, individual symptoms including cough, loss of smell/taste, fever, fatigue, headache, and sore throat were independently associated with generating antibodies following a positive PCR test. The strongest predictors were the four classic symptoms (cough, loss of smell/taste, fever).

We estimated the half-life of anti-spike IgG to be 184 days, indicating a sustained antibody response against infection, compared with previous reports between 36 and 244 days^15,17,19–23^. We found multiple factors associated with peak levels and decline. Variation in the literature may be explained by differences in study design, population (age, sex), and assay performance (different targets and assay types). Longer half-lives were correlated with lower peak levels, suggesting some individuals, e.g. after mild disease^20,27^ mount a lower antibody response that wanes more slowly, whilst others, produce higher antibody responses but that wane more quickly. This contrasts with a previous healthcare worker study which found a positive correlation between IgG half-life and peak levels^15^, but agrees with a study on 963 infected individuals reporting a faster decay of IgG in hospitalized patients with high initial response than individuals with asymptomatic or mild infections^23^. Since most SARS-CoV-2 infection is mild/asymptomatic, the duration of antibody responses in our study are likely to best generalise to the population at large.

As expected from previous studies of humoral immunity, older age was associated with lower seroconversion rates. However, among those that did seroconvert, peak IgG levels were higher in older individuals. Similar findings have been reported in healthcare workers, where older age (in those of working age) was associated with higher maximum anti-nucleocapsid IgG levels and longer half-lives^15^. Others have also reported associations between older age and higher immune responses, including IgG and memory B cells^37^. One postulated mechanism is that older adults exhibit higher IgG levels because they expand their catalogue of memory B and T cells through accumulated memory^38^. However in our study, selection bias may contribute, as our findings are conditional on participants seroconverting, and the subset of older participants who seroconvert may have more robust immune responses than younger participants overall, amongst whom more may seroconvert despite more heterogenous underlying immunity.

Females previously infected with SARS-CoV-2 have been found to have more robust T cell activation and develop stronger antibody responses than males^39,40^. We found that males were equally likely to seroconvert, but among those that did seroconvert, males had a shorter IgG half-life than females, despite no evidence of difference in peak IgG levels, consistent with a previous healthcare worker study^44^. Another study found no difference in IgG antibody between males and females in mild infection and recovering patients, but a higher IgG in females than males in severe infections and early phases of infection^41^. In our study, non-white participants were more likely to seroconvert than white participants (in models not adjusting for Ct value) and to develop higher antibody levels that then waned more quickly. Higher antibody levels in non-white ethnicity have been reported in several healthcare worker populations^15,42^, consistent with our findings. The observed sex and ethnicity effects likely arise from a combination of genetic and societal factors, and studies more fully adjusting for confounding arising from social differences and structural inequalities may be required to estimate the relative contributions of each mechanism.

While lower Ct values were associated with seroconversion, we found that higher Ct values were associated with slightly higher peak IgG levels, which was counterintuitive as higher Ct values (lower viral burden) have been previously associated with lower antibody titres^8,20,43^. The most likely explanation is that as testing was conducted at regular intervals, rather than in response to symptoms, measured Ct values do not fully reflect peak viral load in our study. We found no evidence of association between self-reported symptoms and IgG peak levels or half-life, although symptoms were associated with seroconversion; previous findings suggest that symptomatic infections develop stronger antibody responses than asymptomatic infections^26^. This could be because our models conditioned on those who seroconverted, or because infections in this general population were generally mild.

Important findings from our study are the predictions about the duration of antibody responses associated with protection from infection, albeit that these related to thresholds previously associated with protection from reinfection or protection from severe infection in vaccine trials. Other immune responses may last for differing time periods, and also memory responses may mean that protection lasts longer than measurable antibody levels. Furthermore, we assume that antibody levels fall exponentially; if the rate of decline slows over time, antibody levels may be sustained for longer. We estimated the time from peak level to three thresholds, the positivity threshold 42ng/ml, 28ng/ml (50% protection from any symptomatic/asymptomatic infection^4^), and 6ng/ml (3% of our estimated peak level, providing 50% protection against severe infection according to^24^). Based on extrapolations from other studies correlating anti-spike IgG antibody titres with neutralising activity and early protection (i.e. within a year) from re-infection with currently circulating variants, we found that 50% protection against infection might be expected to last 1.5-2 years, with protection against severe infection lasting several years. However, given that emerging variants may require higher antibody levels for the same level of neutralisation, the duration of protection might be substantially reduced. It may also be the case that the functional quality of antibodies changes over time^44^; this was not evaluated in this study. Overall, at least in the short-term, protection against re-infection appears high.

Study limitations include the fact that we only measured anti-spike IgG using a single assay; seronegative non-responders in Class 3 might have antibodies detected using other assays or other target antigens. We did not measure neutralizing antibodies or T cell responses; however, neutralizing antibody responses are strongly correlated (Spearman ρ=0.87) with anti-spike binding antibodies following infection as previously reported^45^. This community survey had visits scheduled independent of infection or symptom status, so we could not precisely identify the start of infection or symptom onset; we therefore also incorporated positives from the national testing programme (targeting symptomatic infections) and used the first swab positive test and latent class models to indirectly estimate the start of infection. Similarly, we were not able to model antibody trajectories from each participant’s maximum levels since antibody data were collected monthly. However, we chose a starting point that was close to but slightly after the peak IgG level; while this could slightly underestimate peak IgG levels, the half-life will be unbiasedly estimated if the assumption of exponential decline is correct. Re-infections were rare, with only 92 (0.5%) participants with antibody data having potential re-infections >120 days after their first infection episode **(Figure S1)**. Most had only one antibody result, so it was impossible to investigate any boosting of antibody levels following re-infection.

In conclusion, in this representative study of infected individuals from the UK general population, around 1 in 4 people did not develop anti-spike IgG antibodies following a positive-PCR test in regular screening. Non-responders were more likely to be older and not report symptoms. Among participants who seroconvert, anti-spike IgG antibodies remained above the positivity threshold for 347-502 days for 20 year olds, 366-529 days for 40 year olds, 385-552 days for 60 year olds, and 400-571 days for 80 year olds. These estimates of the durability of natural immunity may aid planning of the vaccination strategies. Further studies are required to determine the extent to which waning antibody levels impact immunity and protection following infection and vaccination, and to assess the risk of infection in seronegative non-responders.

## Online Methods

### Population and settings

The United Kingdom’s Office for National Statistics (ONS) COVID-19 Infection Survey (CIS) (ISRCTN21086382) randomly selects private households on a continuous basis from address lists and previous surveys to provide a representative sample across United Kingdom’s four countries (England, Wales, Northern Ireland, Scotland). After obtaining verbal agreement to participate, a study worker visited each household to take written informed consent from individuals ≥2 years. This consent was obtained from parents/carers for those 2-15 years, while those 10-15 years also provided written assent. Children aged <2 years were not eligible for the study.

At the first visit, participants were asked for (optional) consent for follow-up visits every week for the next month, then monthly for 12 months from enrolment. Individuals were surveyed on their socio-demographic characteristics, behaviours, and vaccination status. Combined nose and throat swabs were taken from all consenting household members for SARS-CoV-2 PCR testing.

For a random 10-20% of households, individuals ≥16 years were invited to provide blood samples monthly for serological testing. Participants with a positive swab test and their household members were also invited to provide blood monthly for follow-up visits. Details on the sampling design are provided elsewhere^34^. From April 2021, additional participants were invited to provide blood samples monthly to assess vaccine responses, based on a combination of random selection and prioritisation of those in the study for the longest period (independent of test results). The study protocol is available at https://www.ndm.ox.ac.uk/covid-19/covid-19-infection-survey/protocol-and-information-sheets. The study received ethical approval from the South Central Berkshire B Research Ethics Committee (20/SC/0195).

### Laboratory testing

Combined nose and throat swabs were tested at high-throughput national “Lighthouse” laboratories in Glasgow (from 16 August 2020 to present) and Milton Keynes (from 26 April 2020 to 8 February 2021). The presence of three SARS-CoV-2 genes (ORF1ab, nucleocapsid protein (N), and spike protein (S)) was identified using real-time PCR with the TaqPath RT-PCR COVID-19 kit (Thermo Fisher Scientific). PCR outputs were analysed using UgenTec Fast Finder 3.300.5 (TaqMan 2019-nCoV Assay Kit V2 UK NHS ABI 7500 v2.1; UgenTec), with an assay-specific algorithm and decision mechanism that allows conversion of amplification assay raw data into test results with minimal manual intervention. Samples were called positive if at least a single N and/or ORF1ab gene were detected, and PCR traces exhibited an appropriate morphology. The S gene alone is not considered to be a reliable positive^34^.

SARS-CoV-2 antibody levels were tested on venous or capillary blood samples using an ELISA detecting anti-trimeric spike IgG developed by the University of Oxford^34,46^. Normalised results are reported in ng/ml of mAb45 monoclonal antibody equivalents. Before 26 February 2021, the assay used fluorescence detection as previously described, with a positivity threshold of 8 million units validated on banks of known SARS-CoV-2 positive and negative samples^46^. After this, it used a commercialised CE-marked version of the assay, the Thermo Fisher OmniPATH 384 Combi SARS-CoV-2 IgG ELISA (Thermo Fisher Scientific), with the same antigen and colorimetric detection. mAb45 is the manufacturer-provided monoclonal antibody calibrant for this quantitative assay. To allow conversion of fluorometrically determined values in arbitrary units, we compared 3,840 samples which were run in parallel on both systems. A piece-wise linear regression was used to generate the following conversion formula:

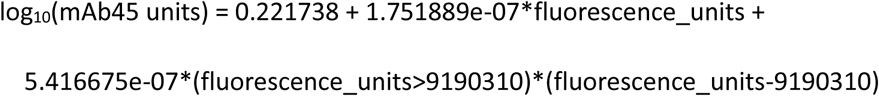

We used 42 ng/ml as the threshold for an IgG positive or negative result (corresponding to the 8 million units with fluorescence detection). We also analysed results using two alternative thresholds, firstly 28 ng/ml (∼7 million fluorescence units), which we had previously found corresponded to 50% protection against any asymptomatic/symptomatic reinfection following a previous infection^4^. We also used 6 ng/ml, the level expected to correspond to 50% protection against severe infection, on the basis of this level of protection being associated with neutralising antibody levels at 3% of peak levels in a previous report^24^. Given the lower and upper limits of the assay, measurements <2 ng/ml (46 observations, 0.3%) and >800 ng/ml (259 observations, 1.8%) were truncated at 2 and 800 ng/ml, respectively.

### Statistical analysis

This analysis included participants aged 16 years and over who had SARS-Cov-2 infection (defined by a positive PCR test) from 26 April 2020 to 14 June 2021. Since multiple positive swab tests could be obtained at follow-up visits, positive PCR tests were grouped into ‘episodes’. We used the first episode (starting with the first positive PCR, or index positive) for each participant in the main analysis. Second episodes, defined by a repeat PCR positive >120 days after the start of the first infection (associated with risk reductions for new positive episodes of similar magnitude to vaccination^47^) were excluded.

Study visits occur on a fixed schedule, meaning that infection episodes could be identified up to 30 days or more after onset (as well as ‘early’ in some pre-symptomatic cases). As participants were told to obtain a test from the national testing programme if symptomatic, to improve our estimate of the start of each infection episode, we linked study data to data on swab positivity from the English national testing programme (data were not available for Scotland, Wales, and Northern Ireland). The national testing programme is intended for individuals with symptoms (although a substantial proportion report no symptoms), and so not all PCR-positive episodes in English study participants also have a positive test from the national testing programme. For this analysis we used the date of the first positive PCR test in the study or the national testing programme as the start of the episode, whichever came first. Ct values and gene positivity patterns are not available from the national testing programme, and so these factors were obtained from PCR-positive samples in the ONS survey only.

We included all antibody measurements from 90 days before each participant’s first swab positive date (index positive) through to 180 days after (approximate 95^th^ percentile), to avoid undue influence from outliers at late time points. We also excluded all antibody measurements taken from 3 days after the first vaccination. Vaccination status was self-reported at study visits, and also linked to the National Immunisation Management Service (NIMS) in England, which contains all individuals’ vaccination data in the English National Health Service COVID-19 vaccination programme. There was good agreement between self-reported and administrative vaccination data (98% on type and 95% on date^47^). We used vaccination data from NIMS where available for participants from England, and otherwise data from the survey.

We used the Ct value as the proxy of viral burden, defined as the minimum from all positive swab tests in the infection episode and categorising at <30 to indicate moderate to higher viral burden. This threshold is used in the UK in algorithms for review of low-level positives at the laboratories where the PCR tests were performed and as a threshold for attempting whole-genome sequencing^47^. Gene positivity pattern during the episode was classified as three groups: 1) a single ORF1ab gene or a single N gene positive; 2) Alpha (B.1.1.7) SARS-CoV-2 variant compatible (at least once positive for ORF1ab+N across the episode and never S positive), and 3) S-positive (ORF1ab+N+S or ORF1ab+S or N+S at least once across the episode). Participants with missing information on Ct values or gene positivity patterns or symptoms in the episode were excluded from analysis (n=133). Self-reported symptoms were those reported at any visit within 35 days after the index positive date or reported to the national testing programme. Fever, cough, loss of smell, and loss of taste were considered ‘classic symptoms’.

We first used latent class mixed models (LCMM) to identify distinct patterns of antibody response after natural infection, counting the date of the index positive in the survey as time 0. Restricted natural cubic splines (internal knots at -10, 30, 60 days, and boundary knots at -60 and 140 days) were used to model time since the index positive as the fixed effect. A random-effect intercept and random-effect slope on all time spline variables were added to account for individual variability. The location of the knots was chosen to reflect fitted antibody trajectories in models with greater numbers of knots, that would not converge while also allowing for random effects. Age as a natural cubic spline (internal knots at 50 years, and boundary knots at 20, 80 years), presence of self-reported long-term health conditions, Ct value, and self-reported symptoms were included as covariates for class membership^48^. The number of classes, up to a maximum of 4, was determined by examining and comparing the shape of the class trajectories and measures of model fit using Bayesian information criterion (BIC).

We used Bayesian linear mixed interval censored models to estimate the decay in antibody responses from their peak level, excluding those who did not seroconvert, and any participant with a positive or equivocal antibody result strictly before their index positive date (≥23 ng/ml) (n=6) or a negative antibody measurement within 42 days of their first index positive (N=13) **(Figure S1)**. Time zero (peak level) for this analysis was determined from the estimated trajectories for each class from the LCMM (see Results). We assumed an exponential fall in antibody levels over time, i.e., a linear decline on a log_2_ scale. Population-level fixed effects, individual-level random effects for intercept and slope, and covariance between random effects were included in the model. The outcome was right-censored at 800 reflecting truncation of IgG values at 800 ng/ml. We excluded a very small number of measurements (24) below 23 ng/ml (likely reflecting mislabelled samples) to reduce the influence of outliers **(Figure S3)**. There was no evidence of non-linearity in antibody decline on the log scale, comparing the main model with a model using natural cubic splines to fit time **(Figure S10**). We also examined the association between peak levels and antibody half-lives with age, sex, ethnicity, reporting having long-term health conditions, Ct values, and self-reported symptoms.

For each Bayesian linear mixed interval censored model, weakly informative priors were used **(Table S7)**. 4 chains were run per model with 4,000 iterations and a warm-up period of 2,000 iterations to ensure convergence, which was confirmed visually and by ensuring the Gelman-Rubin statistic was <1.05 (**Table S5**). 95% credibility intervals were calculated using highest posterior density intervals.

As sensitivity analyses, we additionally used 400 and 500 as the censoring threshold for IgG levels and chose different starting points to examine robustness.

All analyses were performed in R 3.6 using the following packages: tidyverse (version 1.3.0), brms (version 2.14.0), rstanarm (version 2.21.1), splines (version 3.6.1), lcmm (version 1.9.2), nnet (version 7.3-14), ggeffects (version 0.14.3), arsenal (version 3.4.0), cowplot (version 1.1.0), bayesplot (version 1.7.2).

## Supporting information

supplementary results

## Data Availability

Data are still being collected for the COVID-19 Infection Survey. De-identified study data are available for access by accredited researchers in the ONS Secure Research Service (SRS) for accredited research purposes under part 5, chapter 5 of the Digital Economy Act 2017. For further information about accreditation, contact or visit the SRS website.

## Acknowledgements

We are grateful for the support of all COVID-19 Infection Survey participants.

This study is funded by the Department of Health and Social Care with in-kind support from the Welsh Government, the Department of Health on behalf of the Northern Ireland Government and the Scottish Government. ASW, TEAP, NS, DE, KBP are supported by the National Institute for Health Research Health Protection Research Unit (NIHR HPRU) in Healthcare Associated Infections and Antimicrobial Resistance at the University of Oxford in partnership with Public Health England (PHE) (NIHR200915). ASW and TEAP are also supported by the NIHR Oxford Biomedical Research Centre. KBP is also supported by the Huo Family Foundation. ASW is also supported by core support from the Medical Research Council UK to the MRC Clinical Trials Unit [MC_UU_12023/22] and is an NIHR Senior Investigator. PCM is funded by Wellcome (intermediate fellowship, grant ref 110110/Z/15/Z) and holds an NIHR Oxford BRC Senior Fellowship award. DWE is supported by a Robertson Fellowship and an NIHR Oxford BRC Senior Fellowship. NS is an Oxford Martin Fellow and holds an NIHR Oxford BRC Senior Fellowship. The views expressed are those of the authors and not necessarily those of the National Health Service, NIHR, Department of Health, or PHE.

## COVID-19 Infection Survey team group authorship

**Office for National Statistics**: Sir Ian Diamond, Emma Rourke, Ruth Studley, Tina Thomas, Duncan Cook.

**Office for National Statistics COVID Infection Survey Analysis and Operations teams**, in particular Daniel Ayoubkhani, Russell Black, Antonio Felton, Megan Crees, Joel Jones, Lina Lloyd, Esther Sutherland.

**University of Oxford, Nuffield Department of Medicine**: Ann Sarah Walker, Derrick Crook, Philippa C Matthews, Tim Peto, Emma Pritchard, Nicole Stoesser, Karina-Doris Vihta, Jia Wei, Alison Howarth, George Doherty, James Kavanagh, Kevin K Chau, Stephanie B Hatch, Daniel Ebner, Lucas Martins Ferreira, Thomas Christott, Brian D Marsden, Wanwisa Dejnirattisai, Juthathip Mongkolsapaya, Sarah Cameron, Phoebe Tamblin-Hopper, Magda Wolna, Rachael Brown, Sarah Hoosdally, Richard Cornall, David I Stuart, Gavin Screaton.

**University of Oxford, Nuffield Department of Population Health**: Koen Pouwels.

**University of Oxford, Big Data Institute:** David W Eyre, Katrina Lythgoe, David Bonsall, Tanya Golubchik, Helen Fryer.

**University of Oxford, Radcliffe Department of Medicine**: John Bell.

**Oxford University Hospitals NHS Foundation Trust:** Stuart Cox, Kevin Paddon, Tim James.

**University of Manchester**: Thomas House.

**Public Health England**: John Newton, Julie Robotham, Paul Birrell.

**IQVIA**: Helena Jordan, Tim Sheppard, Graham Athey, Dan Moody, Leigh Curry, Pamela Brereton.

**National Biocentre**: Ian Jarvis, Anna Godsmark, George Morris, Bobby Mallick, Phil Eeles.

**Glasgow Lighthouse Laboratory**: Jodie Hay, Harper VanSteenhouse.

**Department of Health**: Jessica Lee.

## Author Contributions

The study was designed and planned by ASW, JF, JB, JN, IB, ID and KBP, and is being conducted by ASW, IB, RS, ER, AH, BM, TEAP, PCM, NS, DA, SH, EYJ, DIS, DWC and DWE. This specific analysis was designed by JW, DWE, ASW, and KBP. JW, KBP and DA contributed to the statistical analysis of the survey data. JW, DWE, KBP and ASW drafted the manuscript and all authors contributed to interpretation of the data and results and revised the manuscript. All authors approved the final version of the manuscript.

## Code availability

A copy of the analysis code is available at https://github.com/jiaweioxford/COVID19_vaccine_antibody_response https://github.com/jiaweioxford/COVID19_infection_antibody_response.

