## supplementary results for "Anti-spike antibody response to natural SARS-CoV-2 infection in the general population"

| | Total (N=7256) | Class 1 'seroconverted' (N=4683) | Class 2 'possible late/reinfection' (N=831) | Class 3 'seronegative non-responders' (N=1742) | p value (all) | P value (2 vs 1) | P value (3 vs 1) |
|---|---|---|---|---|---|---|---|
| **Percentage** | 100% | 64.5% | 11.5% | 24.0% | | | |
| **Symptoms & Ct value** | | | | | < 0.001 | < 0.001 | < 0.001 |
| **No self-reported symptoms & Ct>=30** | 1779 (24.5%) | 132 (2.8%) | 528 (63.5%) | 1119 (64.2%) | | | |
| **No self-reported symptom & Ct<30** | 1287 (17.7%) | 912 (19.5%) | 122 (14.7%) | 253 (14.5%) | | | |
| **Self-reported symptoms & Ct>=30** | 1057 (14.6%) | 641 (13.7%) | 128 (15.4%) | 288 (16.5%) | | | |
| **Self-reported symptom & Ct<30** | 3133 (43.2%) | 2998 (64.0%) | 53 (6.4%) | 82 (4.7%) | | | |
| **Have >=2 positive swabs in the infection episode** | | | | | < 0.001 | < 0.001 | < 0.001 |
| **No** | 4285 (59.1%) | 1975 (42.2%) | 628 (75.6%) | 1682 (96.6%) | | | |
| **Yes** | 2971 (40.9%) | 2708 (57.8%) | 203 (24.4%) | 60 (3.4%) | | | |
| **Days between first and last positive swab if >=2 positive swabs** | | | | | < 0.001 | 0.002 | < 0.001 |
| **Median** | 14 | 14 | 18 | 20 | | | |
| **IQR** | 7, 26 | 7, 26 | 10, 28 | 10, 29 | | | |
| **Have negative study swab before index positive** | | | | | < 0.001 | < 0.001 | < 0.001 |
| **Yes** | 6203 (85.5%) | 4032 (86.1%) | 603 (72.6%) | 1568 (90.0%) | | | |
| **No** | 1053 (14.5%) | 651 (13.9%) | 228 (27.4%) | 174 (10.0%) | | | |
| **Days between previous negative and index positive** | | | | | < 0.001 | < 0.001 | < 0.001 |
| **Median** | 26 | 25 | 29 | 25 | | | |
| **IQR** | 12, 35 | 13, 34 | 25, 60 | 7, 29 | | | |
| **Have national testing programme positive included within infection episode** | | | | | < 0.001 | < 0.001 | < 0.001 |
| **No** | 5675 (78.2%) | 3210 (68.5%) | 756 (91.0%) | 1709 (98.1%) | | | |
| **Yes** | 1581 (21.8%) | 1473 (31.5%) | 75 (9.0%) | 33 (1.9%) | | | |
| **National testing programme positive before study index positive** | | | | | < 0.001 | < 0.001 | < 0.001 |
| **No** | 6053 (83.4%) | 3570 (76.2%) | 764 (91.9%) | 1719 (98.7%) | | | |
| **Yes** | 1203 (16.6%) | 1113 (23.8%) | 67 (8.1%) | 23 (1.3%) | | | |
| **Days between national testing programme positive and study index positive** | | | | | < 0.001 | < 0.001 | < 0.001 |
| **Median** | 7 | 6 | 15 | 16 | | | |
| **IQR** | 3, 14 | 3, 13 | 10, 24 | 10, 23 | | | |

**Table S1. Additional characteristics of classes identified from latent class mixed models for 7,256 participants infected with SARS-CoV-2.** Main characteristics among classes are presented in **Table 1**, and continuous variables are presented graphically in **Figure S2**. Continuous variables were compared using Kruskall-Wallis tests, and categorical variables were compared using Chi-squared tests.

| | Class 1 Seroconverted | Class 2 Possible late/reinfection | | | Class 3 Seronegative | | |
|---|---|---|---|---|---|---|---|
| | | OR | 95%CI | p-value | OR | 95%CI | p-value |
| **Age** | ref | See **Figure 2** | | | | | |
| **Sex** | | | | | | | |
| **Female** | | 1(ref) | | | 1(ref) | | |
| **Male** | | 0.98 | 0.81-1.18 | 0.8 | 0.88 | 0.74-1.04 | 0.1 |
| **Ethnicity** | | | | | | | |
| **White** | | 1(ref) | | | 1(ref) | | |
| **Non-white** | | 1.03 | 0.75-1.4 | 0.9 | 0.8 | 0.6-1.08 | 0.2 |
| **Report having long-term health conditions** | | | | | | | |
| **No** | | 1(ref) | | | 1(ref) | | |
| **Yes** | | 0.82 | 0.64-1.04 | 0.1 | 1.1 | 0.9-1.35 | 0.4 |
| **Report working in patient facing healthcare** | | | | | | | |
| **No** | | 1(ref) | | | 1(ref) | | |
| **Yes** | | 1.02 | 0.52-1.96 | 1 | 0.39 | 0.18-0.83 | **0.014** |
| **Ct value (one unit higher)** | | 1.35 | 1.32-1.39 | **<0.001** | 1.33 | 1.31-1.36 | **<0.001** |
| **Report having symptoms** | | | | | | | |
| **No symptoms** | | 1(ref) | | | 1(ref) | | |
| **Other symptoms** | | 0.09 | 0.07-0.13 | **<0.001** | 0.23 | 0.19-0.29 | **<0.001** |
| **Classic symptoms** | | 0.07 | 0.05-0.09 | **<0.001** | 0.07 | 0.06-0.09 | **<0.001** |
| **Have >=2 positive swabs in the infection episode** | | | | | | | |
| **No** | | 1(ref) | | | 1(ref) | | |
| **Yes** | | 1.13 | 0.8-1.6 | 0.5 | 0.08 | 0.05-0.13 | **<0.001** |
| **Days between first and last positive (1 day longer)** | | 0.99 | 0.98-1 | 0.1 | 1 | 0.98-1.02 | 0.8 |

**Table S2. Odds ratio with 95% confidence intervals from multinomial logistic regression, using Class 1 as the reference.** See **Figure 2** for effect of age and other factors on the probability scale. The 95% confidence intervals are calculated by prediction ± 1.96*standard error of the prediction; Wald p values are shown.

| | Class 1 'seroconverted' | Class 3 (A: Using all data): 'seronegative non responders' | | | Class 3 (B: Conditioning on Ct ≤32 and ≥2 gene positivity) | | |
|---|---|---|---|---|---|---|---|
| | | OR | 95%CI | p-value | OR | 95%CI | p-value |
| **Age** | ref | | | **<0.001** | | | **<0.001** |
| **Sex (Male vs Female)** | | 0.84 | 0.73-0.97 | **0.02** | 0.85 | 0.69-1.04 | 0.1 |
| **Ethnicity (Non-White vs White)** | | 1.26 | 0.98-1.62 | 0.07 | 1.17 | 0.82-1.69 | 0.4 |
| **Report having long-term health conditions (Yes vs No)** | | 1.19 | 1.00-1.43 | 0.05 | 1.06 | 0.81-1.38 | 0.7 |
| **Report working in patient facing healthcare (Yes vs No)** | | 0.42 | 0.21-0.84 | **0.01** | 0.35 | 0.11-1.13 | 0.08 |
| **Cough (Yes vs No)** | | 0.18 | 0.13-0.24 | **<0.001** | 0.20 | 0.13-0.33 | **<0.001** |
| **Loss of smell (Yes vs No)** | | 0.21 | 0.12-0.36 | **<0.001** | 0.31 | 0.15-0.66 | **0.002** |
| **Fever (Yes vs No)** | | 0.38 | 0.24-0.58 | **<0.001** | 0.40 | 0.21-0.77 | **0.006** |
| **Loss of taste (Yes vs No)** | | 0.45 | 0.28-0.72 | **<0.001** | 0.47 | 0.23-0.95 | **0.03** |
| **Fatigue/weakness (Yes vs No)** | | 0.58 | 0.43-0.80 | **<0.001** | 0.70 | 0.44-1.11 | 0.1 |
| **Headache (Yes vs No)** | | 0.64 | 0.48-0.86 | **0.003** | 0.60 | 0.39-0.94 | **0.03** |
| **Sore throat (Yes vs No)** | | 0.63 | 0.46-0.88 | **0.006** | 0.55 | 0.32-0.92 | **0.02** |
| **Myalgia (Yes vs No)** | | 0.83 | 0.59-1.18 | 0.3 | 0.66 | 0.38-1.15 | 0.1 |
| **Nausea/Vomiting (Yes vs No)** | | 0.84 | 0.50-1.43 | 0.5 | 1.11 | 0.52-2.35 | 0.8 |
| **Diarrhoea (Yes vs No)** | | 0.95 | 0.53-1.72 | 0.9 | 0.74 | 0.27-2.01 | 0.6 |
| **Abdominal pain (Yes vs No)** | | 0.96 | 0.50-1.83 | 0.9 | 0.96 | 0.35-2.63 | 0.9 |
| **Shortness of breath (Yes vs No)** | | 1.39 | 0.93-2.10 | 0.1 | 1.39 | 0.76-2.53 | 0.3 |
| **Angina (Yes vs No)** | | 0.96 | 0.43-2.14 | 0.9 | 1.04 | 0.38-2.82 | 0.9 |
| **Asthma (Yes vs No)** | | 0.84 | 0.66-1.06 | 0.1 | 0.93 | 0.67-1.30 | 0.7 |
| **Atrial fibrillation (Yes vs No)** | | 1.07 | 0.59-1.94 | 0.8 | 1.04 | 0.46-2.35 | 0.9 |
| **Cancer (Yes vs No)** | | 1.07 | 0.80-1.43 | 0.6 | 1.03 | 0.69-1.56 | 0.9 |
| **Chronic kidney disease (Yes vs No)** | | 1.13 | 0.70-1.84 | 0.6 | 0.94 | 0.46-1.94 | 0.9 |
| **Chronic liver disease (Yes vs No)** | | 0.88 | 0.39-1.99 | 0.8 | 0.92 | 0.29-2.94 | 0.9 |
| **COPD (Yes vs No)** | | 0.69 | 0.40-1.17 | 0.2 | 1.37 | 0.72-2.60 | 0.3 |
| **Coronary heart disease (Yes vs No)** | | 0.99 | 0.40-2.43 | 1 | 1.19 | 0.39-3.66 | 0.8 |
| **Frailty (Yes vs No)** | | 0.64 | 0.41-1.01 | 0.05 | 0.48 | 0.24-0.98 | **0.04** |
| **Heart failure (Yes vs No)** | | 0.97 | 0.37-2.60 | 1 | 0.93 | 0.26-3.35 | 0.9 |
| **Hypertension (Yes vs No)** | | 0.87 | 0.64-1.17 | 0.3 | 0.81 | 0.52-1.24 | 0.3 |
| **Myocardial infarction (Yes vs No)** | | 1.45 | 0.61-3.48 | 0.4 | 1.21 | 0.37-3.98 | 0.8 |

| | | | | | | |
|---|---|---|---|---|---|---|
| **Osteoporosis (Yes vs No)** | 2.00 | 1.07-3.73 | **0.03** | 1.33 | 0.52-3.42 | 0.6 |
| **Peripheral arterial disease (Yes vs No)** | 0.66 | 0.32-1.37 | 0.3 | 0.63 | 0.22-1.81 | 0.4 |
| **Rheumatoid arthritis (Yes vs No)** | 0.82 | 0.32-2.12 | 0.7 | 0.89 | 0.23-3.53 | 0.9 |
| **Stroke (Yes vs No)** | 1.11 | 0.47-2.62 | 0.8 | 0.53 | 0.11-2.59 | 0.4 |
| **Transient ischaemic attack (Yes vs No)** | 0.77 | 0.32-1.84 | 0.6 | 1.50 | 0.50-4.50 | 0.5 |
| **Type 1 diabetes (Yes vs No)** | 3.06 | 0.95-9.85 | 0.06 | 3.10 | 0.52-18.55 | 0.2 |
| **Type 2 diabetes (Yes vs No)** | 1.29 | 0.76-2.18 | 0.3 | 0.94 | 0.43-2.05 | 0.9 |
| **Overweight (BMI 25 to <30 kg/m$^2$) (Yes vs No)** | 0.73 | 0.61-0.88 | **<0.001** | 0.80 | 0.61-1.05 | 0.1 |
| **Obese (BMI ≥30 kg/m$^2$) (Yes vs No)** | 0.84 | 0.68-1.03 | 0.1 | 0.88 | 0.65-1.19 | 0.4 |
| **Antihypertensive medication (Yes vs No)** | 1.00 | 0.76-1.33 | 1 | 0.95 | 0.64-1.42 | 0.8 |
| **Diabetes medication (Yes vs No)** | 0.47 | 0.26-0.86 | **0.01** | 0.45 | 0.17-1.15 | 0.09 |
| **Corticosteroids (Yes vs No)** | 1.53 | 0.87-2.71 | 0.1 | 0.83 | 0.34-2.04 | 0.7 |
| **Immunosuppressants (Yes vs No)** | 0.79 | 0.31-2.05 | 0.6 | 0.96 | 0.24-3.81 | 1 |

**Table S3. Odds ratio with 95% confidence intervals from a logistic regression model comparing seronegative vs seroconverting participants (Class 3 vs Class 1) using demographic factors, individual symptoms, and comorbidities.** (A) Using all data from Class 3 (N=1,383) vs Class 1 (N=4,032) (B) Restricting Class 3 to those with Ct value ≤32 and ≥2 genes detected (N=487) to decrease the impact of potential false positive swab tests. Age was fitted using natural cubic spline with one internal knot placed at 50 years and two boundary knots at 20, 80 years. Comorbidities were obtained by linkage to the General Practice Extraction Service (GPES) Data for Pandemic Planning and Research and Hospital Episode Statistics (HES) via the NHS number. The 95% confidence intervals are calculated by prediction ± 1.96*standard error of the prediction; Wald p values are shown.

| | | Peak level (ng/ml) | | | Half-life (days) | | |
|---|---|---|---|---|---|---|---|
| | | Posterior mean | 95% Crl | | Posterior mean | 95% Crl | |
| **Model from 56 days after index positive** | Censored at 800 (base case) | 203 | 190 | 210 | 184 | 163 | 210 |
| | Censored at 500 | 194 | 189 | 200 | 187 | 168 | 209 |
| | Censored at 400 | 189 | 183 | 194 | 192 | 172 | 215 |
| **Model from 28 days after index positive** | Censored at 800 | 225 | 213 | 232 | 189 | 171 | 211 |
| | Censored at 500 | 216 | 210 | 222 | 184 | 168 | 202 |
| | Censored at 400 | 209 | 204 | 215 | 187 | 172 | 204 |
| **Model from 84 days after index positive** | Censored at 800 | 177 | 164 | 184 | 225 | 182 | 287 |
| | Censored at 500 | 168 | 163 | 175 | 233 | 192 | 290 |
| | Censored at 400 | 164 | 158 | 170 | 242 | 199 | 305 |

**Table S4. Sensitivity analysis using different starting point (28, 84 days) and different censoring threshold (400, 500 ng/ml) for anti-spike IgG trajectory modelling (peak and half-life estimation).**

| | | Estimate | Est.Error | 95%Crl | | Rhat | Bulk_ESS | Tail_ESS |
|---|---|---|---|---|---|---|---|---|
| **Baseline** | Intercept | 7.6636 | 0.0247 | 7.6149 | 7.7119 | 1.0020 | 2802 | 4046 |
| | time | -0.0054 | 0.0003 | -0.0061 | -0.0048 | 1.0004 | 8113 | 6588 |
| | sigma | 0.5165 | 0.0117 | 0.4943 | 0.5401 | 1.0045 | 261 | 856 |
| | sd(Intercept) | 1.0406 | 0.0250 | 0.9925 | 1.0895 | 1.0010 | 651 | 1953 |
| | sd(time) | 0.0043 | 0.0009 | 0.0021 | 0.0058 | 1.0087 | 137 | 211 |
| | cor(Intercept,time) | -0.2576 | 0.0976 | -0.4166 | -0.0467 | 1.0015 | 1365 | 1047 |
| **Age** | Intercept | 7.6405 | 0.0244 | 7.5932 | 7.6881 | 1.0016 | 2299 | 4221 |
| | time | -0.0049 | 0.0004 | -0.0057 | -0.0042 | 1.0006 | 6927 | 6056 |
| | age | 0.1194 | 0.0163 | 0.0871 | 0.1509 | 1.0044 | 1988 | 3551 |
| | time:age | -0.0001 | 0.0002 | -0.0005 | 0.0004 | 1.0015 | 5853 | 6699 |
| | sigma | 0.5164 | 0.0117 | 0.4935 | 0.5398 | 1.0151 | 304 | 798 |
| | sd(Intercept) | 1.0222 | 0.0239 | 0.9762 | 1.0695 | 1.0078 | 645 | 2515 |
| | sd(time) | 0.0043 | 0.0009 | 0.0023 | 0.0058 | 1.0355 | 147 | 186 |
| | cor(Intercept,time) | -0.2409 | 0.0943 | -0.4008 | -0.0338 | 1.0057 | 920 | 906 |
| **Sex** | Intercept | 7.6376 | 0.0339 | 7.5700 | 7.7040 | 1.0012 | 2330 | 4247 |
| | time | -0.0044 | 0.0005 | -0.0053 | -0.0034 | 1.0005 | 5928 | 6931 |
| | sex | 0.0531 | 0.0488 | -0.0429 | 0.1486 | 1.0007 | 2225 | 3689 |
| | time:sex | -0.0022 | 0.0007 | -0.0035 | -0.0009 | 1.0005 | 6714 | 6566 |
| | sigma | 0.5179 | 0.0118 | 0.4954 | 0.5417 | 1.0218 | 224 | 421 |
| | sd(Intercept) | 1.0393 | 0.0246 | 0.9910 | 1.0879 | 1.0076 | 575 | 2035 |
| | sd(time) | 0.0041 | 0.0010 | 0.0015 | 0.0057 | 1.0448 | 119 | 113 |
| | cor(Intercept,time) | -0.2615 | 0.1057 | -0.4333 | -0.0319 | 1.0069 | 1667 | 1136 |
| **Ethnicity** | Intercept | 7.6216 | 0.0259 | 7.5706 | 7.6722 | 1.0019 | 2110 | 3747 |
| | time | -0.0053 | 0.0004 | -0.0060 | -0.0046 | 1.0008 | 5942 | 6477 |
| | ethnicity | 0.4344 | 0.0833 | 0.2660 | 0.5929 | 1.0005 | 2263 | 4012 |
| | time:ethnicity | -0.0018 | 0.0011 | -0.0039 | 0.0004 | 1.0001 | 5954 | 6204 |
| | sigma | 0.5163 | 0.0115 | 0.4937 | 0.5392 | 1.0052 | 267 | 1149 |
| | sd(Intercept) | 1.0340 | 0.0245 | 0.9880 | 1.0831 | 1.0038 | 626 | 2504 |
| | sd(time) | 0.0043 | 0.0009 | 0.0024 | 0.0059 | 1.0131 | 113 | 334 |
| | cor(Intercept,time) | -0.2512 | 0.0927 | -0.4074 | -0.0427 | 1.0033 | 953 | 1222 |
| **LTHC** | Intercept | 7.6294 | 0.0276 | 7.5750 | 7.6833 | 1.0011 | 2125 | 4128 |
| | time | -0.0054 | 0.0004 | -0.0062 | -0.0047 | 1.0004 | 7285 | 6868 |
| | LTHC | 0.1646 | 0.0630 | 0.0416 | 0.2879 | 1.0006 | 2121 | 3843 |
| | time:LTHC | 0.0002 | 0.0009 | -0.0017 | 0.0020 | 1.0007 | 5814 | 6214 |
| | sigma | 0.5188 | 0.0119 | 0.4959 | 0.5429 | 1.0308 | 213 | 568 |
| | sd(Intercept) | 1.0347 | 0.0248 | 0.9870 | 1.0828 | 1.0145 | 484 | 842 |
| | sd(time) | 0.0040 | 0.0010 | 0.0014 | 0.0057 | 1.0409 | 122 | 238 |
| | cor(Intercept,time) | -0.2406 | 0.1093 | -0.4122 | 0.0049 | 1.0076 | 1159 | 747 |
| **Ct value** | Intercept | 7.6610 | 0.0247 | 7.6127 | 7.7098 | 1.0008 | 2161 | 4383 |
| | time | -0.0055 | 0.0004 | -0.0062 | -0.0048 | 1.0001 | 7177 | 6326 |
| | ct | 0.0056 | 0.0040 | -0.0024 | 0.0133 | 1.0015 | 2173 | 4200 |
| | time:ct | 0.0000 | 0.0001 | -0.0001 | 0.0001 | 1.0002 | 6296 | 7132 |
| | sigma | 0.5170 | 0.0117 | 0.4953 | 0.5414 | 1.0088 | 217 | 293 |
| | sd(Intercept) | 1.0390 | 0.0244 | 0.9914 | 1.0869 | 1.0044 | 530 | 1126 |
| | sd(time) | 0.0042 | 0.0010 | 0.0016 | 0.0058 | 1.0200 | 108 | 168 |

| | | | | | | | | |
|---|---|---|---|---|---|---|---|---|
| | cor(Intercept,time) | -0.2540 | 0.1041 | -0.4155 | -0.0414 | 1.0059 | 947 | 507 |
| **Symptom** | Intercept | 7.6801 | 0.0543 | 7.5737 | 7.7884 | 1.0004 | 2481 | 4294 |
| | time | -0.0066 | 0.0008 | -0.0081 | -0.0051 | 1.0002 | 6162 | 5701 |
| | symptom | -0.0195 | 0.0608 | -0.1395 | 0.0996 | 1.0012 | 2486 | 4307 |
| | time:symptom | 0.0014 | 0.0009 | -0.0002 | 0.0031 | 1.0002 | 5915 | 6392 |
| | sigma | 0.5157 | 0.0126 | 0.4922 | 0.5411 | 1.0200 | 223 | 475 |
| | sd(Intercept) | 1.0407 | 0.0256 | 0.9900 | 1.0907 | 1.0118 | 428 | 1126 |
| | sd(time) | 0.0043 | 0.0010 | 0.0019 | 0.0060 | 1.0330 | 117 | 155 |
| | cor(Intercept,time) | -0.2565 | 0.0982 | -0.4161 | -0.0412 | 1.0071 | 845 | 818 |
| **Multivariable** | Intercept | 7.5307 | 0.0603 | 7.4120 | 7.6482 | 1.0022 | 2235 | 3459 |
| | time | -0.0045 | 0.0009 | -0.0062 | -0.0027 | 1.0003 | 6706 | 5743 |
| | age | 0.1336 | 0.0169 | 0.1011 | 0.1672 | 1.0001 | 2283 | 4077 |
| | sex | 0.0464 | 0.0488 | -0.0510 | 0.1416 | 1.0009 | 2368 | 4226 |
| | ethnicity | 0.5297 | 0.0817 | 0.3720 | 0.6927 | 1.0006 | 2408 | 3995 |
| | LTHC | 0.0994 | 0.0613 | -0.0178 | 0.2175 | 1.0009 | 2373 | 4184 |
| | ct | 0.0091 | 0.0040 | 0.0012 | 0.0168 | 1.0000 | 2178 | 3914 |
| | symptom | 0.0088 | 0.0593 | -0.1065 | 0.1269 | 1.0028 | 2029 | 4134 |
| | time:age | -0.0002 | 0.0002 | -0.0007 | 0.0003 | 1.0001 | 6309 | 6334 |
| | time:sex | -0.0021 | 0.0007 | -0.0034 | -0.0008 | 1.0002 | 7569 | 6581 |
| | time:ethnicity | -0.0021 | 0.0011 | -0.0042 | 0.0000 | 1.0003 | 6654 | 7010 |
| | time:LTHC | 0.0000 | 0.0009 | -0.0018 | 0.0018 | 1.0004 | 7363 | 7377 |
| | time:ct | 0.0000 | 0.0001 | -0.0001 | 0.0001 | 0.9999 | 7213 | 6771 |
| | time:symptom | 0.0009 | 0.0009 | -0.0007 | 0.0027 | 1.0002 | 5384 | 6424 |
| | sigma | 0.5186 | 0.0122 | 0.4950 | 0.5431 | 1.0040 | 224 | 406 |
| | sd(Intercept) | 1.0064 | 0.0252 | 0.9576 | 1.0563 | 1.0038 | 385 | 931 |
| | sd(time) | 0.0038 | 0.0012 | 0.0008 | 0.0056 | 1.0160 | 131 | 171 |
| | cor(Intercept,time) | -0.2162 | 0.1272 | -0.4037 | 0.0933 | 1.0030 | 586 | 326 |

**Table S5. Model coefficients and MCMC diagnostics for the univariable and multivariable models.** The reference category in the multivariable model is: 43-year-old, female, white ethnicity, no long-term health conditions, Ct value=22, and no self-reported symptoms.

| A | | 42 ng/ml | | 28 ng/ml | | 6 ng/ml | |
|---|---|---|---|---|---|---|---|
| **Age** | **Sex&ethnicity** | **Estimate** | **95%CrI** | **Estimate** | **95%CrI** | **Estimate** | **95%CrI** |
| **20** | White female | 502 | 239-980 | 644 | 364-1232 | 1186 | 782-2051 |
| | Non-white female | 436 | 261-747 | 530 | 341-898 | 887 | 621-1470 |
| | White male | 361 | 192-580 | 454 | 280-698 | 815 | 590-1180 |
| | Non-white male | 347 | 220-526 | 417 | 281-619 | 686 | 504-980 |
| **40** | White female | 529 | 289-926 | 660 | 409-1117 | 1163 | 802-1909 |
| | Non-white female | 457 | 284-748 | 547 | 364-875 | 885 | 631-1406 |
| | White male | 386 | 228-590 | 476 | 308-697 | 817 | 609-1122 |
| | Non-white male | 366 | 240-536 | 433 | 299-627 | 693 | 522-972 |
| **60** | White female | 552 | 316-989 | 673 | 418-1196 | 1140 | 781-1966 |
| | Non-white female | 476 | 306-798 | 562 | 378-917 | 884 | 628-1448 |
| | White male | 408 | 251-623 | 495 | 330-737 | 819 | 605-1158 |
| | Non-white male | 385 | 258-565 | 448 | 317-651 | 699 | 525-1000 |
| **80** | White female | 571 | 335-1180 | 686 | 422-1385 | 1119 | 733-2056 |
| | Non-white female | 495 | 320-896 | 574 | 380-1023 | 882 | 613-1605 |
| | White male | 430 | 272-692 | 511 | 341-817 | 821 | 587-1276 |
| | Non-white male | 400 | 275-618 | 464 | 327-712 | 704 | 515-1075 |
| **B** | | 42 ng/ml | | 28 ng/ml | | 6 ng/ml | |
| **Group** | **Multiplier** | Estimate | 95%CrI | Estimate | 95%CrI | Estimate | 95%CrI |
| **60-year-old white male** | 1 | 409 | 251-626 | 495 | 330-732 | 819 | 605-1156 |
| | 2 | 261 | 112-441 | 347 | 195-552 | 673 | 486-970 |
| | 3 | 178 | 57-341 | 261 | 112-441 | 588 | 412-853 |
| | 5 | 68 | 57-222 | 155 | 57-323 | 479 | 312-714 |
| | 10 | 58 | 57-75 | 58 | 57-161 | 333 | 183-530 |
| **60-year-old white female** | 1 | 551 | 317-990 | 674 | 425-1184 | 1138 | 776-1955 |
| | 2 | 344 | 131-660 | 463 | 241-842 | 930 | 625-1596 |
| | 3 | 218 | 57-486 | 344 | 131-660 | 809 | 525-1406 |
| | 5 | 64 | 57-305 | 186 | 57-450 | 653 | 404-1154 |
| | 10 | 58 | 57-69 | 58 | 57-196 | 444 | 224-826 |

**Table S6. A) Posterior predicted days with 95% credibility interval from the start of infection to three anti-spike IgG thresholds (42, 28, and 6 ng/ml) by age (20, 40, 60, 80 years), sex, and ethnicity in the multivariable model. B) Posterior predicted days with 95% credibility interval from the start of infection to thresholds (42, 28, and 6 ng/ml) multiplied by 2, 3, 5, 10 in 60-year-old white male and female population.**

| Model term | Priors |
|---|---|
| Intercept | normal (7.4, 2.8) |
| Slope | normal (0, 0.1) |
| Coefficient for change in intercept (age) | normal (0, 2) |
| Coefficient for change in slope (age) | normal (0, 0.1) |
| Coefficient for change in intercept (sex) | normal (0, 6) |
| Coefficient for change in slope (sex) | normal (0, 0.1) |
| Coefficient for change in intercept (ethnicity) | normal (0, 10) |
| Coefficient for change in slope (ethnicity) | normal (0, 0.2) |
| Coefficient for change in intercept (long-term health condition) | normal (0, 7) |
| Coefficient for change in slope (long-term health condition) | normal (0, 0.2) |
| Coefficient for change in intercept (Ct) | normal (0, 1) |
| Coefficient for change in slope (Ct) | normal (0, 0.1) |
| Coefficient for change in intercept (symptom) | normal (0, 6) |
| Coefficient for change in slope (symptom) | normal (0, 0.1) |
| Random effect SD: intercept | normal (0, 1) |
| Random effect SD: slope | normal (0, 0.01) |
| Random effect intercept & slope covariance | lkj_corr_cholesky (2) |

**Table S7. Priors used in the Bayesian linear mixed interval censored models.**

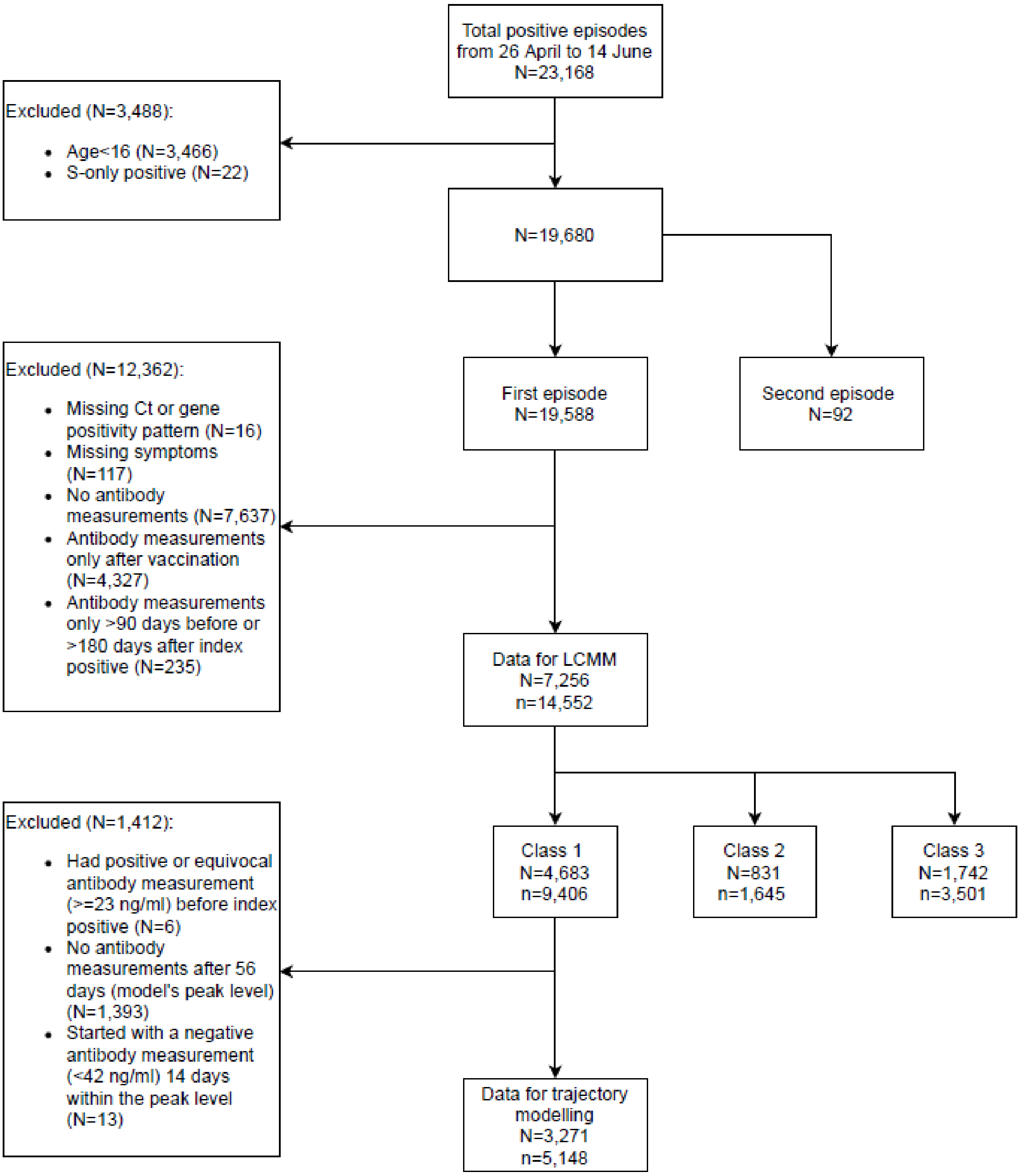


**Figure S1. Flowchart of the study cohort.** N represents the number of positive episodes/participants, and n represents the number of antibody measurements. LCMM=latent class mixed model. Class 1='seroconverted in response to infection', Class 2='possible late/reinfection', Class 3='seronegative non-responders', see **Figure 1** and **Figure S3**.

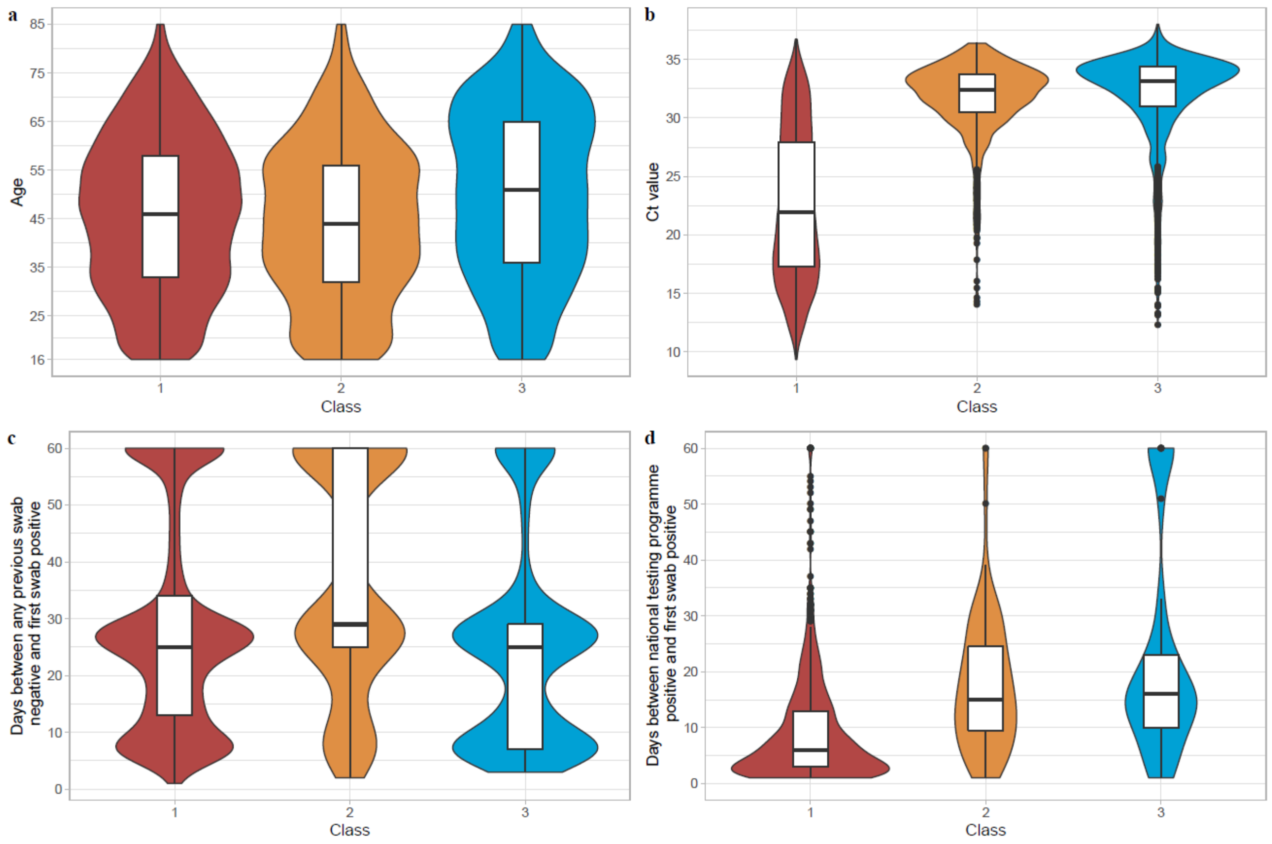


**Figure S2. Distributions of continuous variables by classes identified from latent class mixed models. a**, Participants' age. Participants in Class 3 were older than Class 1 and 2. **b**, Participants' minimum Ct value across the infection episode. Class 1 had a lower Ct value than Class 2 and 3. **c**, Days between any previous study swab negative test and the first swab positive test in the infection episode. More participants in Class 2 had a longer duration between previous negative swab and the index positive, supporting late detection. **d,** Days between any preceding national testing programme positive test and the first swab positive test (y-axis truncated at 60 days for visualisation). Class 1 had a shorter duration between the two tests than other two classes. Class 1='seroconverted in response to infection' (N=4,683, 64.5%), Class 2='possible late/reinfection' (N=831, 11.5%), Class 3='seronegative non-responders' (N=1,742, 24.0%). For the box and whisker inserts: centre line, median; box limits, upper and lower quartiles; whiskers, 1.5x interquartile range; points, outliers.

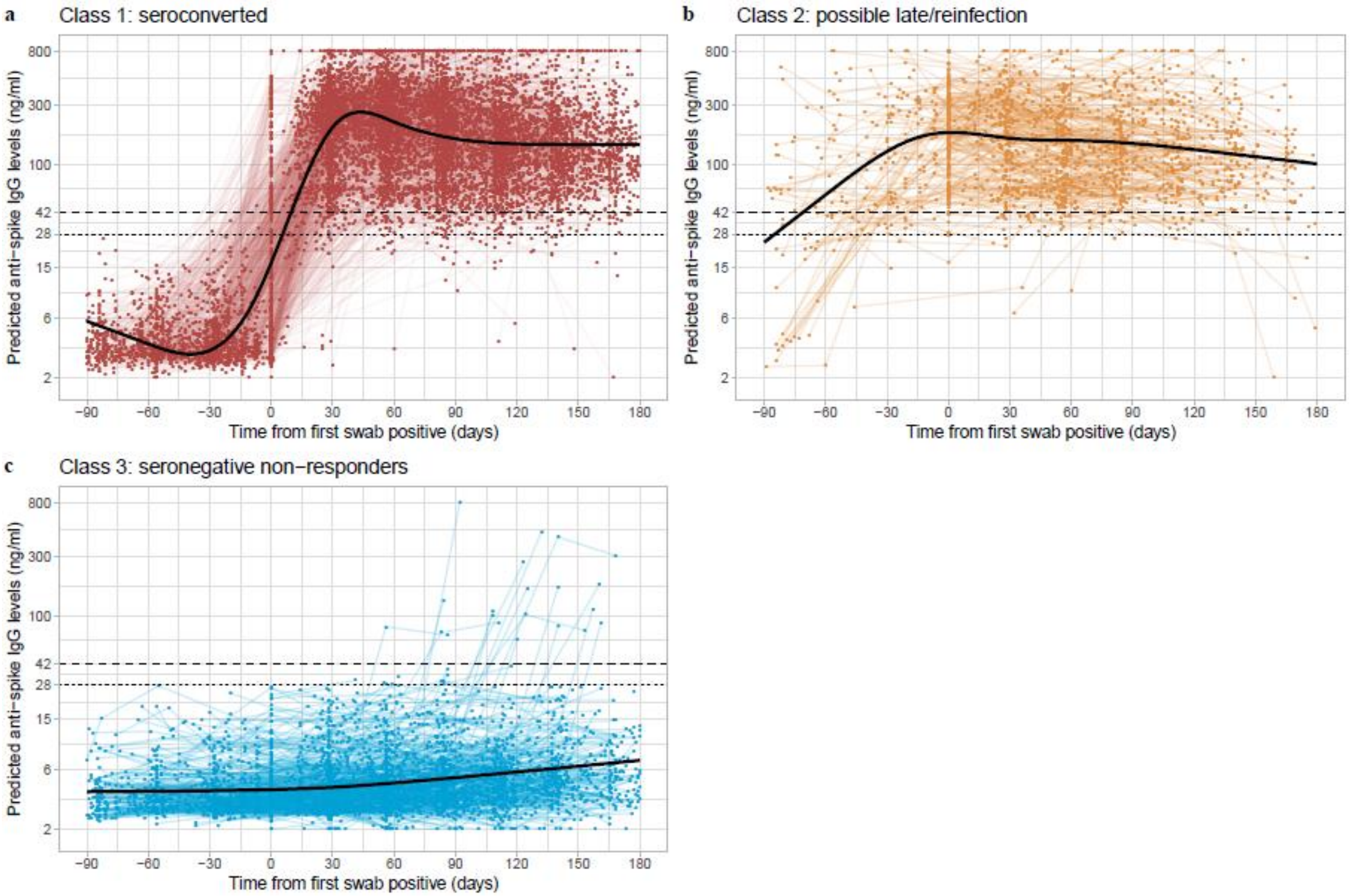


**Figure S3. Individual trajectories for 7,256 participants infected with SARS-CoV-2 by class identified from latent class mixed models**. **a,** Class 1, 'seroconverted in response to infection' (N=4,683, 64.5%). **b,** Class 2, 'possibly late/reinfection' (N=831, 11.5%). **c,** Class 3, 'seronegative non-responders' (N=1,742, 24.0%). Black dashed line indicates the assay threshold for IgG positivity (42 ng/ml) and the dotted line at 28 ng/ml indicates level associated with 50% protection against reinfection.

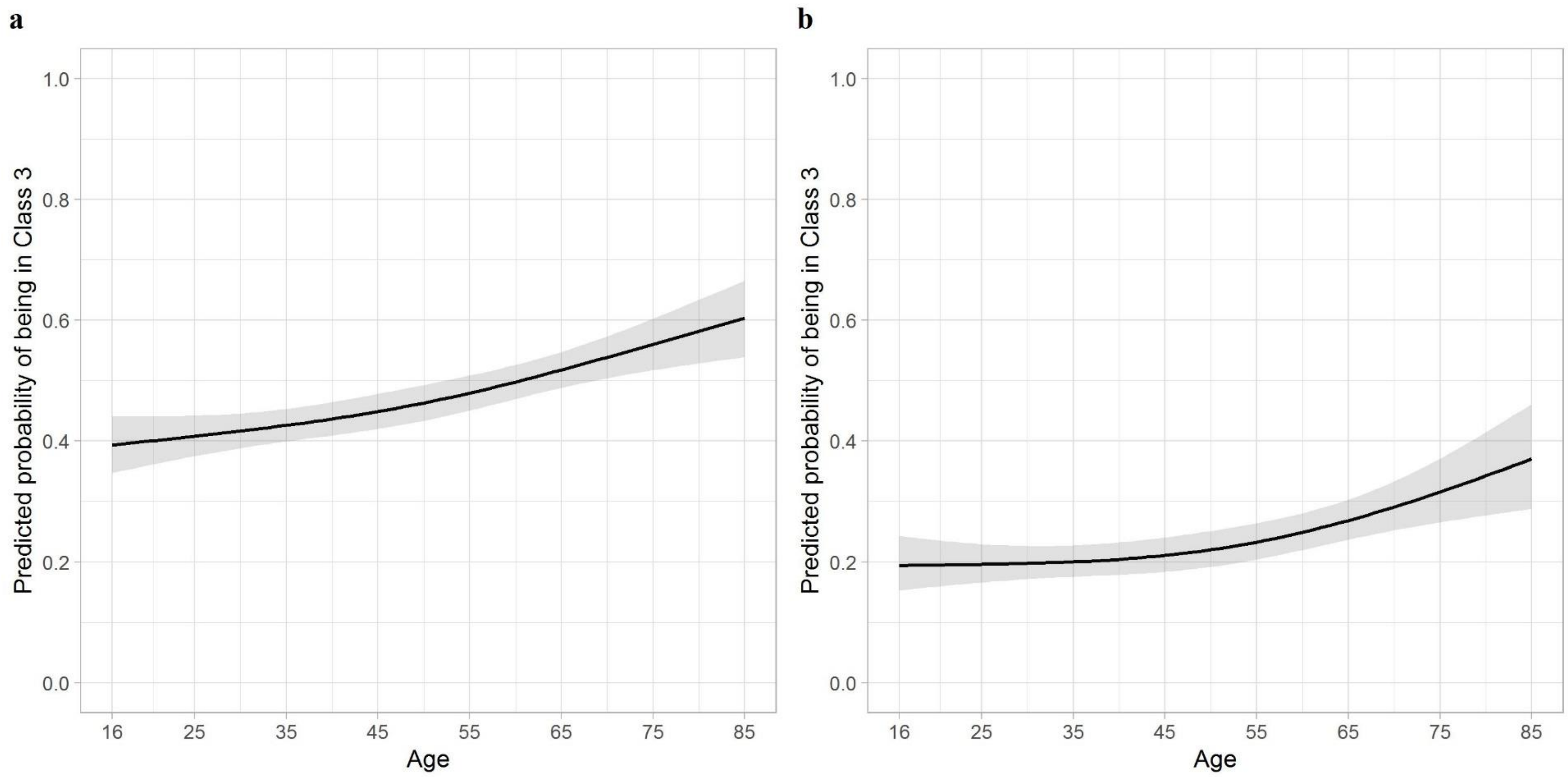


**Figure S4. Predicted probability of being in Class 3 (seronegative) compared with Class 1 (seroconverted) by age.** Plotted at the reference category for other variables (female, white ethnicity, no long-term health condition, not working in patient-facing healthcare, no self-reported symptoms of any kind)**.** **a,** Using all data from Class 3 (N=1,742). **b**, Conditioning on Ct value ≤32 and ≥2 genes detected (N=595) to decrease the impact of potential false positive swab tests. Age was fitted using natural cubic spline with one internal knot placed at 50 years and two boundary knots at 20, 80 years. Full model's results are presented in Table 2. The 95% confidence intervals are calculated by prediction ± 1.96*standard error of the prediction.

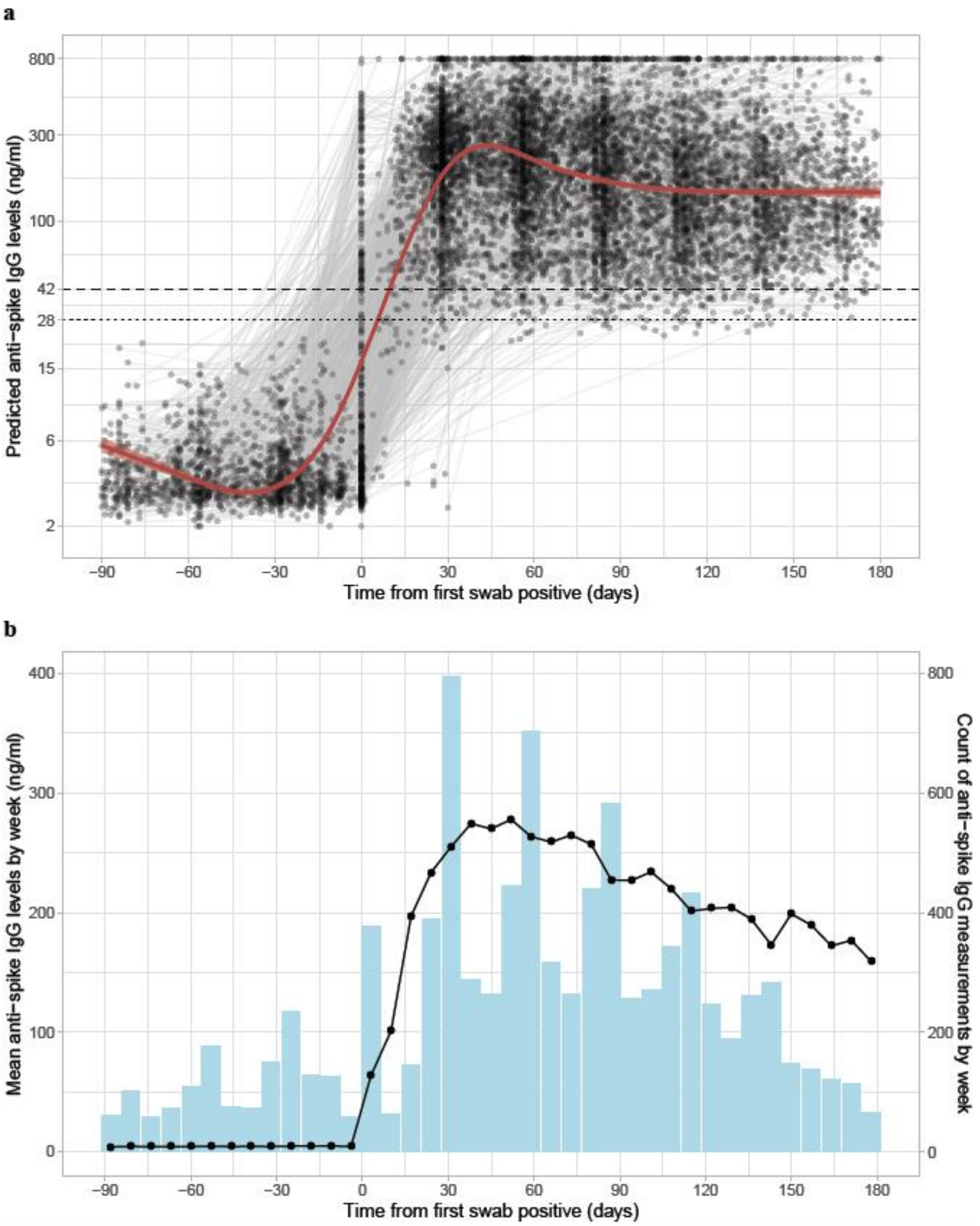


**Figure S5**. **Determination of the 'peak level' for modelling anti-spike IgG trajectories. a,** Class trajectory and individual trajectories for participants included in the model from Class 1. **b,** Mean anti-spike IgG levels and count of IgG measurements by week after index positive test. Around 56 days (8 weeks), the number of IgG measurements is high, and the mean IgG levels are close to the highest level.

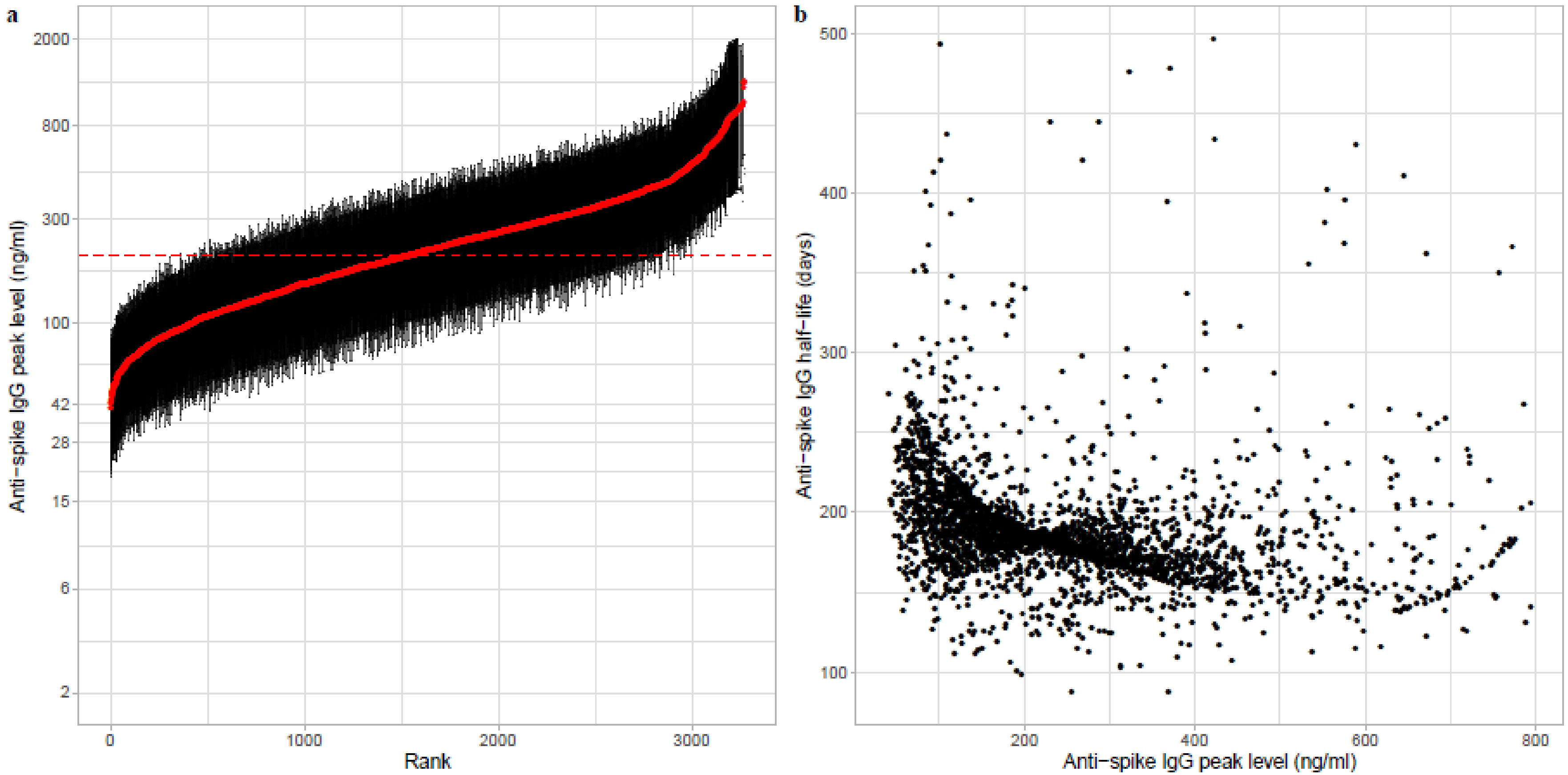


**Figure S6. Posterior estimations for individual anti-spike IgG half-lives and peak level. a,** Estimated peak anti-spike IgG level with 95% CrI for all participants (N=3,271) by rank. The red dot shows the mean level in each individual. The red dashed line shows the overall mean level (203 ng/ml) . **b,** Relationship between estimated IgG peak levels with half-lives. X and Y axes are truncated at 800 and 500 for visualization (Spearman's rank coefficient=-0.5, p<0.0001).

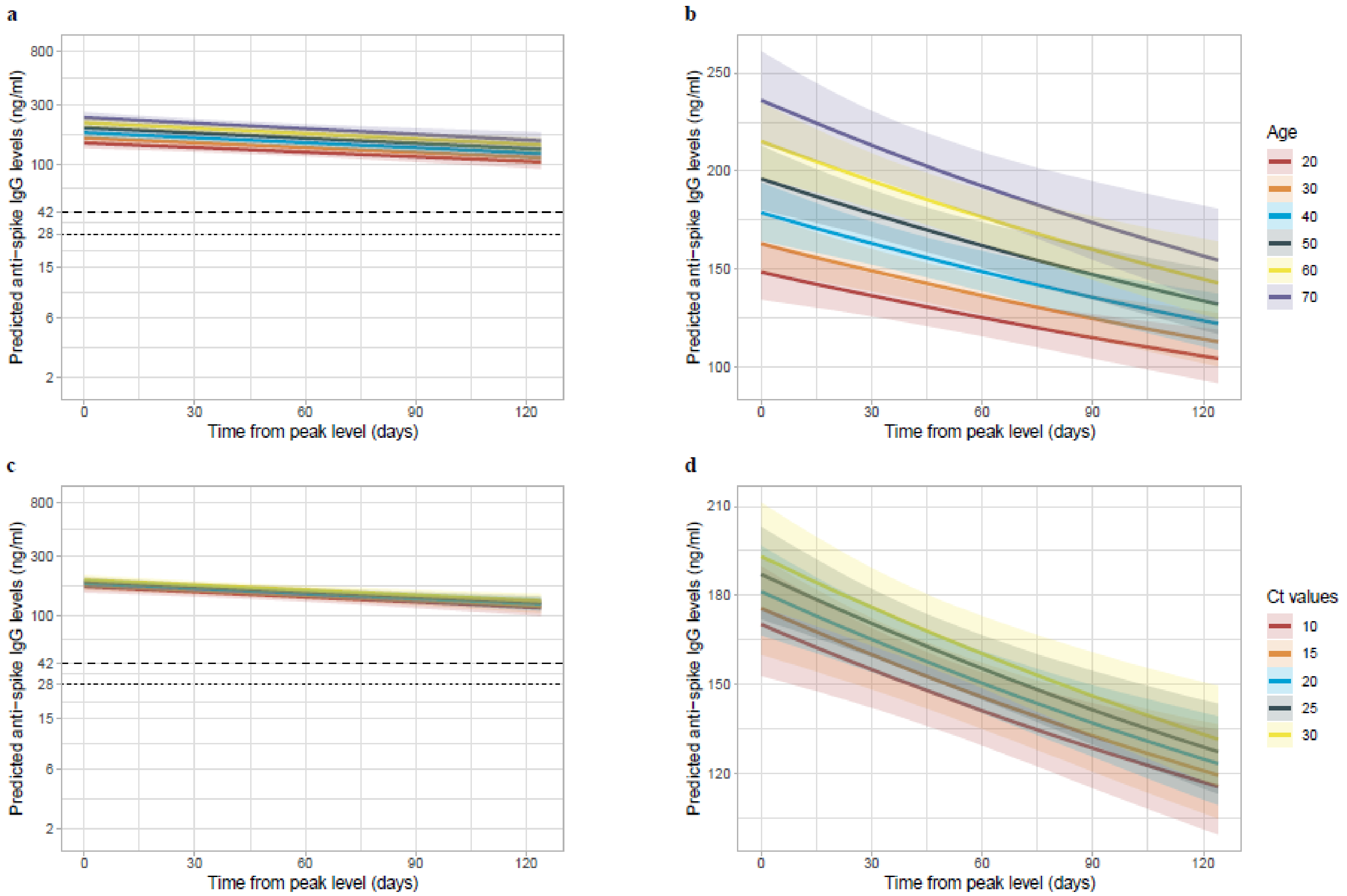


**Figure S7. Posterior mean trajectories of anti-spike IgG after the peak level by age (a, b) and Ct values (c, d) in the multivariable model in 3,271 participants.** Panel a and c show the relationship on the log10 scale. Black dashed line indicates the assay threshold for IgG positivity (42 ng/ml) and the dotted line at 28 ng/ml (indicates level associated with 50 % protection against reinfection). Panel b and d show the relationship in its original scale. Full model results are shown in Table 3.

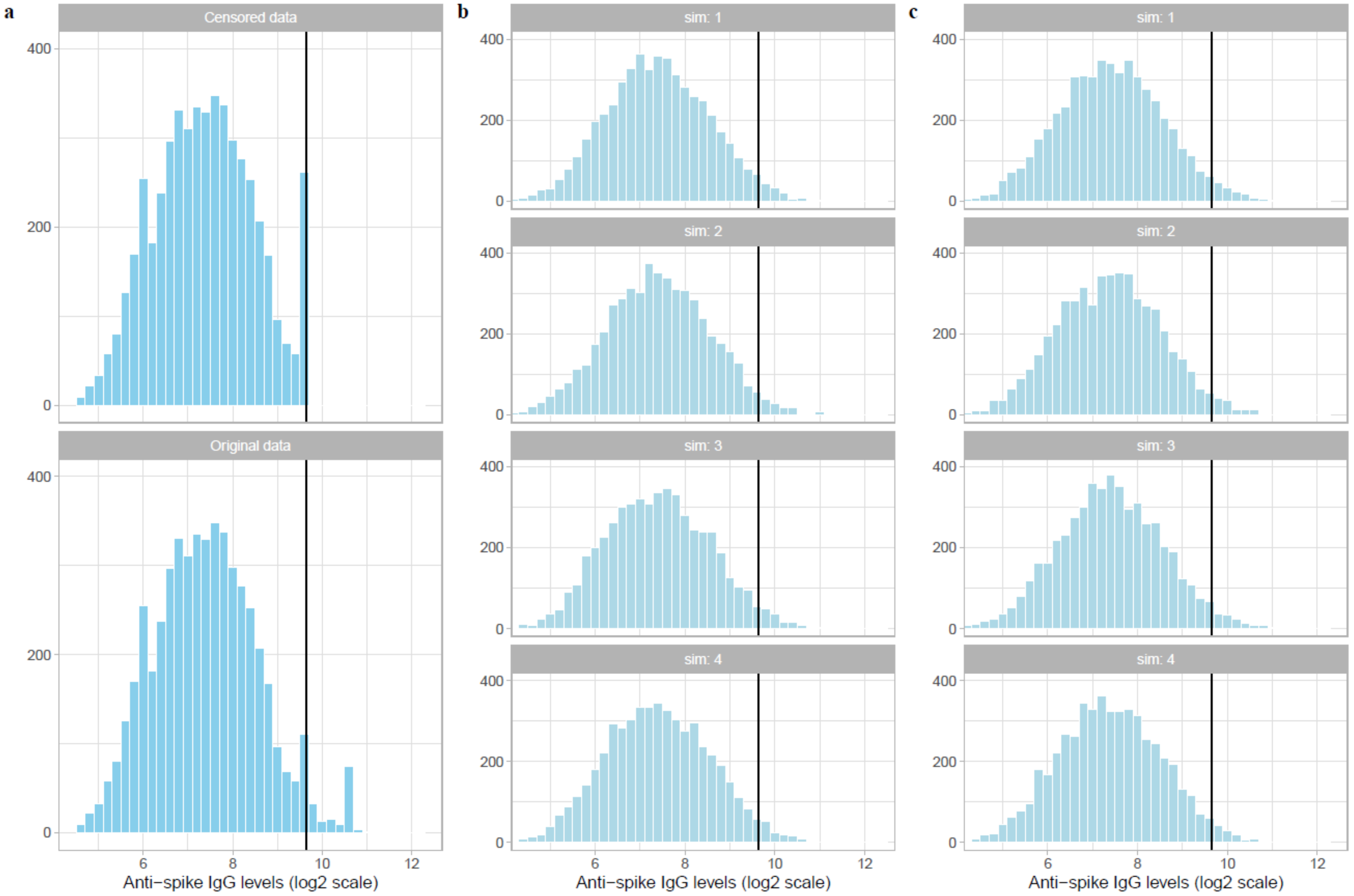


**Figure S8. Posterior predictive check for the baseline model and the multivariable model. a,** Distribution of the observed anti-spike IgG levels on log2 scale. **b,** Distribution of anti-spike IgG levels from four posterior simulated datasets for the baseline model. **c,** Distribution of anti-spike IgG levels from four posterior simulated datasets for the multivariable model.

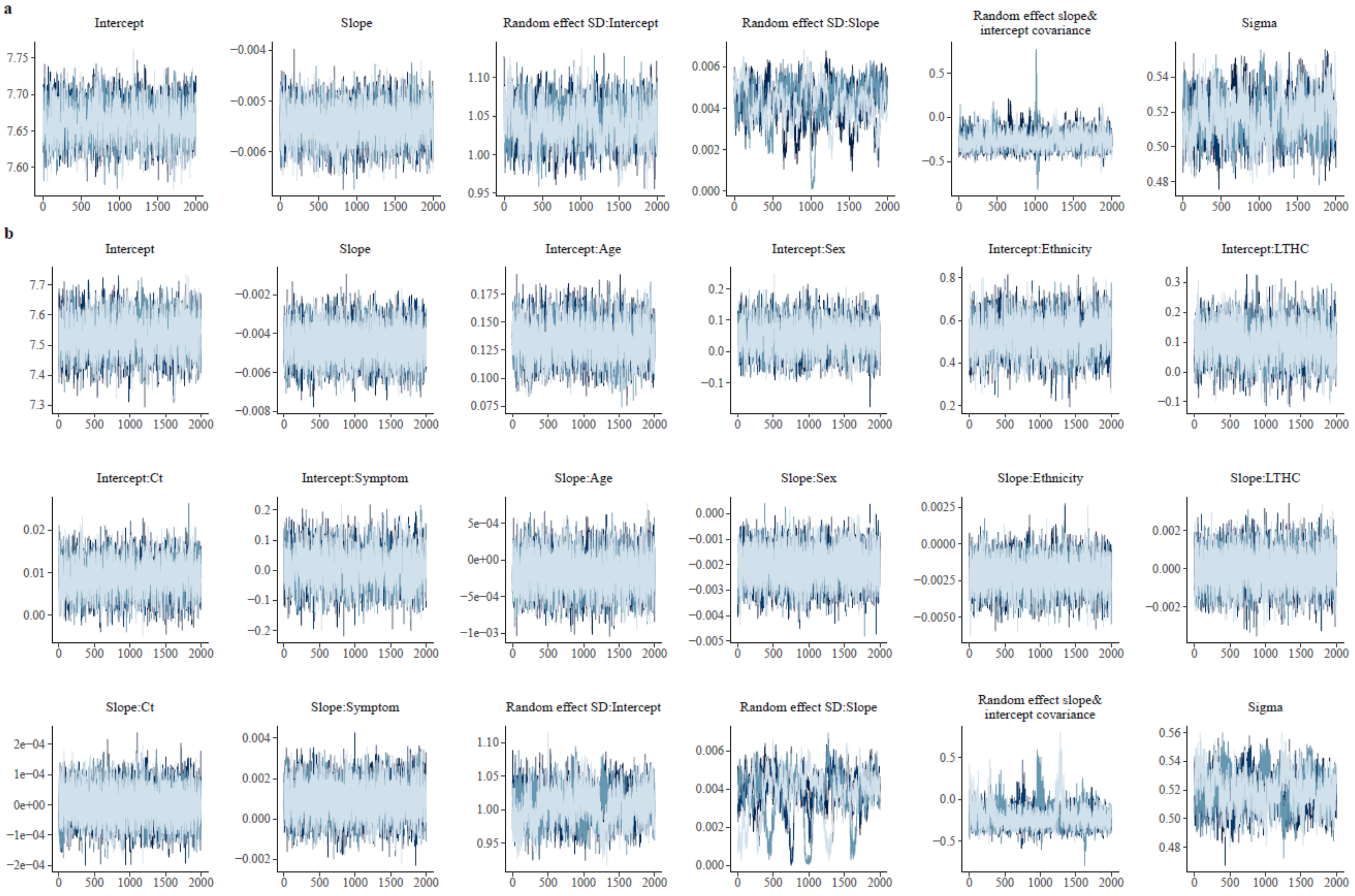


**Figure S9. MCMC trace plots for assessing convergence of chains. a,** Trace plots for the baseline model. **b,** Trace plots for the multinomial model.

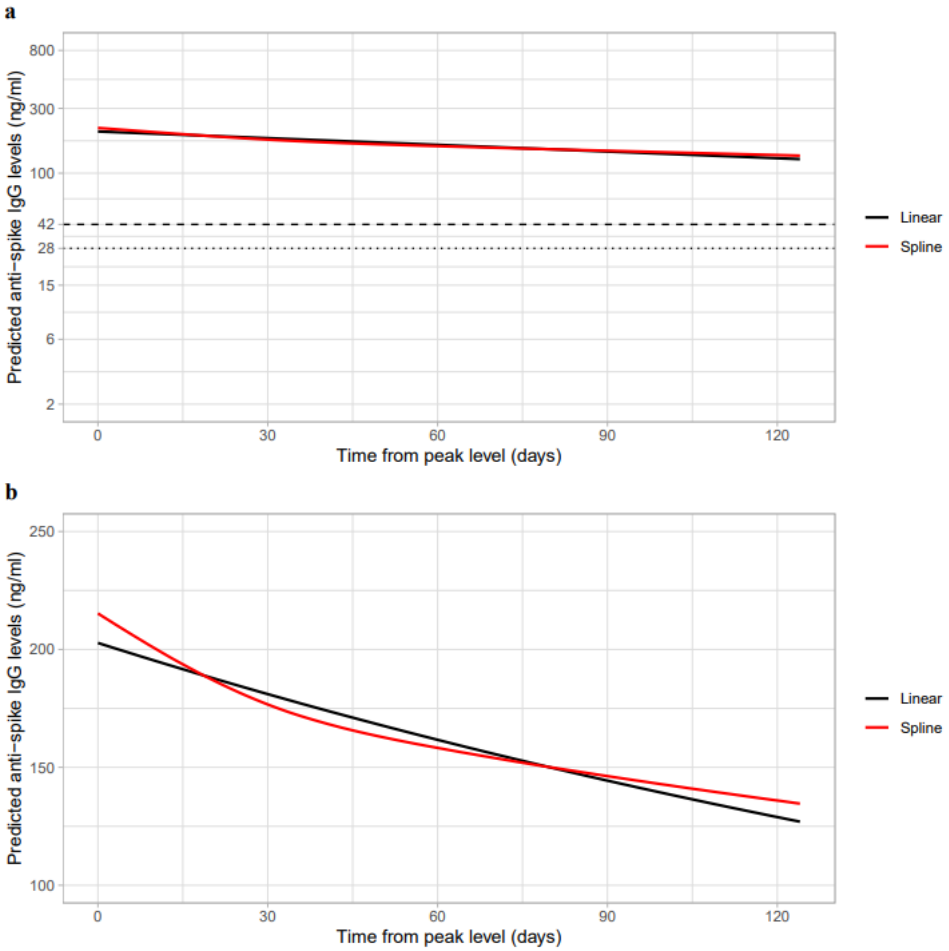


**Figure S10. Comparison of anti-spike IgG decline in 3,271 participants between the model assuming a linear decline in log2 scale (black line) and the model using splines (red line).** Time was fitted using natural cubic splines with internal knots at 30, 70 and boundary knots at 5, 110. Panel a shows the trajectory on the log10 scale. Black dashed line indicates the assay threshold for IgG positivity (42 ng/ml) and the dotted line at 28 ng/ml (indicates level associated with 50 % protection against reinfection). Panel b shows the trajectory in its original scale. There is no evidence of non-linearity in antibody decline.
